# Research and Diagnostic Algorithmic Rules (RADAR) for mood disorders, recurrence of illness, suicidal behaviors, and the patient’s lifetime trajectory

**DOI:** 10.1101/2022.09.15.22279968

**Authors:** Michael Maes, Juliana Brum Moraes, Ana Congio, Heber Odebrecht Vargas, Sandra Odebrecht Vargas Nunes

**Author notes:** Corresponding author: Prof. Dr. Michael Maes, M.D., Ph.D. Department of Psychiatry, Faculty of Medicine Chulalongkorn University, Bangkok, 10330, Thailand, Michael Maes - Google Scholar.

## Abstract

The top-down DSM/ICD categories of mood disorders are inaccurate, and their dogmatic nature precludes both deductive (as indisputable) and inductive (as top-down) remodeling of case definitions. In trials, psychiatric rating scale scores employed as outcome variables are invalid and rely on folk psychology-like narratives. Using machine learning techniques we developed a new precision nomothetic model of mood disorders with a recurrence of illness (ROI) index, a new endophenotype class, namely Major Dysmood Disorder (MDMD), characterized by increased ROI, a more severe phenome, and more disabilities Nonetheless, our previous studies did not compute Research and Diagnostic Algorithmic Rules (RADAR) to diagnose MDMD and score ROI, lifetime (LT), and current suicidal behaviors, as well as the phenome of mood disorders. Here we provide rules to compute bottom-up RADAR scores for MDMD, ROI, lifetime (LT) and current suicidal SI and SA, the phenome of mood disorders, and the lifetime trajectory of mood disorder patients from a family history of mood disorders and substance abuse to adverse childhood experiences, ROI, and the phenome. We also demonstrate how to plot the 12 major scores in a single RADAR graph, which displays all features in a two-dimensional plot. These graphs allow the characteristics of a patient to be displayed as an idiomatic fingerprint, allowing one to estimate the key traits and severity of the illness at a glance. Consequently, biomarker research into mood disorders should use our RADAR scores to examine pan-omics data, which should be used to enlarge our precision models and RADAR graph.

## 1. Introduction

Recently, we reviewed that there are no correct (reliable and verified) models of major depression (MDD), a major depressive episode (MDE) and bipolar disorder (BD) (Maes, 2022; Maes and Stoyanov, 2022). Not only is there no correct model of clinical depression, but there is an assortment of various conceptual frameworks, classifications, model conceptions, including a non-model, for the same condition (Maes, 2022). When diagnosing depression, it is as though psychiatrists cannot comprehend one another and speak different languages including psychoanalysis, folk psychology, psychodynamic psychiatry, systemic therapy, self-system theory, cognitive-behavioral therapy, and mind-brain dualism, and other non-falsifiable theories (Maes and Stoyanov, 2022). More evidence of the tremendous chaos that characterizes the current approach of depression is the multitude of depression subtypes, subclasses, and labels (atypical depression, melancholia, recurrent depressive disorder, dysthymia, bipolar depression, double depression, psychotic depression, persistent depressive disorder, treatment resistant depression) along with the ongoing formation of new depression categories with new case definitions that may disappear after a time (reactive depression, situational depression, vital depression, endogenous depression, endogenomorph depression, hidden depression, concealed depression, anxious depression, and a mixed episode in bipolar disorders) (Maes and Stoyanov, 2022). As such, mood disorders research is beset with extreme noise leading to a complete cacophony of terms and labels without a solid consensus among psychiatrists.

In addition, there are no models that distinguish MDD from common emotional distress responses including loss, grieving, and demoralization (Maes, 2022). Folk psychologists and sociologists appear to believe that clinical depression is a boundary experience or that “psychiatry transformed normal sorrow into depressive disorder,” contributing to the medicalization of normal human behavior such as feeling blue, grieving, sadness, and demoralization (van Os and Kohne, 201; Kohne and Van Os, 2021; Summerfield, 2006; Frances, 2013). As a result, a serious medical sickness such as recurrent major depression is often regarded a boundary experience, and in psychiatric research, severe medical phenotypes are lumped together with common emotional distress responses (Maes, 2022).

The Western (and thus worldwide) gold standard is to diagnose mood disorders using DSM (American Psychiatric Association, 2013) or ICD (World Health Organization, 2004) criteria, which take into account diverse subtypes such as MDD, MDE and BD, either type 1 (manic) or type 2 (hypomanic) episodes. However, DSM/ICD definitions of mood disorders are unreliable, invalid, and often lack consensus among psychiatrists, resulting in a high incidence of misdiagnosis and misclassification whereby the same subtype may be underrated or overrated (Maes, 2022; Maes and Stoyanov, 2022). Not only are the DSM/ICD classifications of mood disorders inaccurate, but their dogmatic character prohibits deductive (as incontestable) and inductive (as top-down) remodeling of the case-definitions (Maes, 2022). In actuality, psychiatric diagnoses in general are inaccurate post-hoc, higher-order constructs based on clinical narratives of the condition (Maes, 2022). As such, their usage as explanatory variable is not only a conceptual error, but also leads to a vast number of errors and invalid conclusions (Maes, 2022). Given that machine learning permits the construction and cross-validation of new correct phenotypes and endophenotype classes (Maes and Stoyanov, 2022), it is incomprehensible that psychiatric research continues to use invalid outcome measurements, such as the DSM/ICD classifications, as an explanatory variable in research papers (Maes, 2022).

Recently, we developed a new clinimetrics method based on supervised and unsupervised machine learning to construct new (pathway) phenotypes and (endo)phenotype classes, namely precision nomothetic psychiatry (Maes et al., 2021a; Maes, 2022; Simeonova et al., 2021; Maes et al., 2022c; Stoyanov and Maes, 2021). Nomothetic refers to the tendency to derive mathematical laws (rules) from indicator (independent) variables that explain the variability in a phenomenon thereby permitting model generalization and providing a covering law model for the illness (Hyman, 2010; Stoyanov, 2020; Maes, 2022; Stoyanov and Maes, 2021). The precision nomothetic models and theories of mood disorders that we constructed encompass the relevant building blocks of an illness, namely the causome (genetic load and adverse childhood experiences (ACE) or their interactions), adverse outcome pathways (pathways and biomarkers), the recurrence of illness (ROI, including episodes and suicidal behaviors), the phenome of depression, consisting of the symptomatome (severity of depression and anxiety, current suicidal behaviors and cognitive deficits), and the phenomenome (the self-experience of illness as assessed by using self-rated health-related quality of life (HR-QoL) and disability scores) (Maes, 2022).

Machine learning enabled us to construct a) novel data-driven nomothetic models of mood disorders, b) a new pathway phenotype using factor analysis, namely a ROI index based on the recurrence of episodes and suicide attempts (SA), and c) new endophenotype class using cluster analysis, namely Major Dysphoric Disorder (MDMD) versus simple DMD (SDMD) (Maes et al., 2021a; Maes, 2022; Simeonova et al., 2021; Maes et al., 2022c; Stoyanov and Maes, 2021). MDMD is characterized by more severe symptoms, more suicidal behaviors, higher ROI, greater cognitive impairments, activated immunological and nitro-oxidative stress pathways, decreased HR-QOL, and increased disability (Maes et al., 2021a; Maes, 2022; Simeonova et al., 2021; Maes et al., 2022c). As such, we constructed covering law models of MDMD, a serious medical disorder characterized by neuro-immune and neuro-oxidative pathways.

However, our published precision nomothetic algorithms, which include neuro-immune and neuro-oxidative stress pathways, are not applicable to other scientific centers and medical practice because such biomarker assays are not standardized throughout research centers and cannot always be evaluated. In addition, our prior studies did not intend to compute novel Research and Diagnostic Algorithmic Rules (RADAR) useful in research settings and clinical practice to diagnose MDMD, and evaluate ROI, lifetime (LT) and current suicidal ideation (SI) and (SA) attempts, and the phenome (symptomatome + phenomenome) of mood disorders. Moreover, our previous models and theories did not differentiate between LT and current SI and SA, nor did we examine the effects of a positive family history of psychiatric disorders on the defining characteristics of mood disorders.

Hence, the present study was conducted to develop new RADAR scores that are applicable in scientific research and clinical practice and permit the measurement of MDMD, ROI, suicidal behaviors (SB), the phenome, and the lifetime course of mood disorders. The second objective is to display a unique fingerprint of the patient’s condition by summarizing all features of the disease in RADAR scores that are displayed in RADAR (or star or spider) plots.

## Methods

### Participants

Patients were referred from the University of the State of Londrina (UEL), Parana, Brazil, and were treated as outpatients at UEL’s University Hospital. Subjects serving as controls were solicited by word of mouth from residents of the study region. The index episode was not of hypomanic or manic polarity, and all individuals with affective disorders were either in full or partial remission. We included men and women between 18 and 70 years old, and all self-identified races and ethnicities were included. The Diagnostic and Statistical Manual of Mental Disorders, Fourth Edition, Text Revision (DSM-IV-TR) was used to make diagnoses of MDD, MDE, BD (types 1 and 2), anxiety disorders, namely panic disorder, post-traumatic stress disorder (PTSD), obsessive-compulsive disorder (OCD), generalized anxiety disorder (GAD), social phobia and specific phobia, and tobacco use disorder (TUD). Participants with a) immune and autoimmune disorders, such as cancer, psoriasis, inflammatory bowel disease, chronic kidney disease, type 1 diabetes, rheumatoid arthritis, and Chrohn’s disease; b) neuroinflammatory and neurodegenerative diseases, such as multiple sclerosis, Alzheimer’s disease, Parkinson’s disease, and stroke; c) other axis-1 DSM-IV-TR disorders, including autism, schizophrenia, schizo-affective disorder, substance use (except tobacco use) and psycho-organic disorders were excluded. Normal controls were additionally excluded for any psychiatric axis-1 disorder.

In all, 172 people were included in the study: 67 healthy controls and 105 people with affective disorders, divided into 68 people with BD, and 37 people with MDD. In addition, 84 people met the criteria for one of the anxiety disorders recognized by the DSM-IV-TR, including panic disorder (n=27), OCD (n=12), PTSD (n=7), GAD (n=46), social phobia (n=14), and specific phobia (n=39).

The study was approved by the UEL Research Ethics Committee (protocol number: CAAE 34935814.2.0000.5231). All participants gave written informed consent prior to taking part in the study.

### Assessments

Using a validated Portuguese version of the structured clinical interview for DSM-IV axis I (Del-Ben et al., 2001) and DSM-TR diagnostic criteria, mood disorder patients were divided into MDD, and BD, type 1 and type 2. Through a semi-structured interview, the psychiatrist conducting the study collected demographic (marital, educational, financial, and occupational) and clinical (number of previous depressive, hypomanic, and manic episodes) information. This rater also scored the familial load in first degree relatives of TUD (FHIS2), alcohol use disorder (FHIS3), substance (illicit drugs) use disorders (SUD, FHIS4), MDD (FHIS5), BD (FHIS6), SA (FHIS7), and schizophrenia (FHIS8). ACEs were measured using the Childhood Trauma Questionnaire (CTQ), a self-rated scale assessing sexual abuse, physical abuse, emotional abuse, emotional neglect, and physical neglect (Bernstein et al., 2003). In our study, we used a validated Brazilian Portuguese translation (Grassi-Oliviera et al., 2006). Lifetime (LT) SI and SA and current SI and SA were evaluated using a validated Portuguese version of the Columbia-Suicide Severity Rating Scale (C-SSRS) (Posner et al., 2011). We not only rated current (last month) SI/SA but also the last 5 years C-SSRS SA items (5Y SA). Only one patient scored positive on current (last month) C-SSRS SA and, therefore, these data could not be used in the analyses. The severity of depressive and anxious mood was assessed using the Hamilton Depression (HAMD) and Anxiety (HAMA) Rating Scales (Hamilton, 1959; 1960), whilst the overall degree of illness, was measured using the Clinical Global Impression (CGI) scale (Guy, 1976). The Sheehan scale (Sheehan et al., 1996) is a self-reported rating scale that includes questions on daily functioning in areas including work and school (item 1), social life and recreation (item 2), family life and household responsibilities (item 3), days lost last week (item 4) and days unproductive last week (item 5). HR-QoL was assessed using a validated Brazilian Portuguese version of the World Health Organization Quality of Life Instrument-Abbreviated Form (WHOQOL-BREF) (Skevington et al., 2004; Fleck et al., 2000). Physical health (domain 1), mental health (domain 2), social relationships (domain 3), and the environment (domain 4) are the four dimensions of HR-QoL that are probed by WHO-QoL-BREF. Another psychiatrist (AC), blind to the clinical data, assessed the Verbal Fluency Test (VFT), letter and animal category; the Trail-Making Test (TMT), parts A and B, and the Stroop color and word test (SCWT) (part I: neutral trial that measures reaction times, part II: congruent trial, part III: incongruent trial) (Stroop, 1935; Jensen, 1965; Reitan, 1955; Troyer et al., 1997).

We determined body mass index (BMI) by dividing kilogram weight by meter squared (in m2). The International Diabetes Federation’s diagnostic criteria were used to establish a diagnosis of metabolic syndrome (MetS) (Alberti et al., 2006). According to the reported usage of psychotropic medications, some patients were prescribed antidepressants (n=48), lithium (n=26), atypical antipsychotics (n=32), and anticonvulsant mood stabilizers (n=33). In this study we examined whether the diverse constructs were affected by use of these psychotropic drugs.

### Statistics

We used analysis of variance (ANOVA) to examine differences in continuous variables between groups and analysis of contingency tables (χ^2^-test) to examine relationships between nominal variables. Pearson’s product moment and Spearman’s rank order correlation coefficients were used to examine correlations between scale variables and the point-biserial correlation coefficients to check the association between binary and scale variables. The associations between clinical scores and diagnostic classes were examined using univariate GLM analyses, while adjusting for possible confounders such as age, sex, and education. Multiple regression analysis was employed to examine the effects of explanatory variables (e.g. ACEs, sex, a positive family history) on dependent variables (e.g. ROI and phenome scores). We additionally employed a forward stepwise automatic regression technique with p-values of 0.05 (to-enter) and 0.06 (to remove) for inclusion and exclusion in the final regression model.

We produced unstandardized B and/or the standardized β coefficients with t-statistics and exact p-values for each of the explanatory variables in the final regression models, as well as F statistics (and p values) and total variance (R^2^ or partial eta squared, also used as effect size) explained by the model. Collinearity and multicollinearity were examined utilizing tolerance (cut-off value of <0.25), the variance inflation factor (cutoff value >4), and the condition index and variance proportions from the collinearity diagnostics table. The White and modified Breusch-Pagan tests were utilized to confirm the existence of heteroskedasticity. All above tests were two-tailed, and an alpha value of 0.05 was deemed statistically significant. We performed dimension reduction using principal component (PC) or factor analysis (unweighted least squares) using SPSS version 28 to examine whether the 5 ACE, 8 FHIS or LT/current SI/SA data could be reduced to meaningful PCs/factors. To be accepted as a valid PC/factor, the first factor should explain >50% of the variance in the data, all loadings on this factor should be > 0.6 and the KMO should be > 0.7. A visual binning method was applied to split latent variable scores into mutually exclusive classes of individuals. All analyses were conducted using version 28 of the IMB-SPSS program for Windows.

Partial least squares (PLS) path analysis (Ringle et al., 2014) was used as a predictive modeling method that allows one to predict the final outcome (output) variables, namely the phenome of mood disorders using a set of independent (input) variables. **Figure 1** presents the conceptual framework or theory employed in the current study. The phenome of mood disorders was conceptualized as a latent vector extracted from current depression, anxiety, suicidal behavior, HR-QoL and disability data (Maes et al., 2021a). The predictors were ACE and FHIS data. Using PLS, we determined the effects of ACE and FHIS data being mediated via a ROI indicator, which was conceptualized as a factor extracted from number of depression and (hypo)mania episodes (ROI1 index). Furthermore, we constructed separate models to delineate whether the effects of ACE/FHIS on current suicidal behaviors were mediated via LT SI/SA and ROI. Toward this end, we extracted meaningful factors from all LT SI and SA data of the C-SSRS and from number of depression, (hypo)mania and total number of episodes. The next step was to extract one valid factor from the number of depressive and hypomanic episodes, and LT SI/SA data (preferably reduced to some factors) which constitute a ROI2 index. Consequently, we tried to extract one factor from the VFT, TMT and SCWT test scores to delineate whether these data could be reduced into one meaningful z-unit-based composite score. The final step was to construct a model based on ACES/FHIS data, ROI, cognitive deficits, and the phenome of mood disorders. In addition, we included gender, age, and education as input variables.

**Figure 1.**
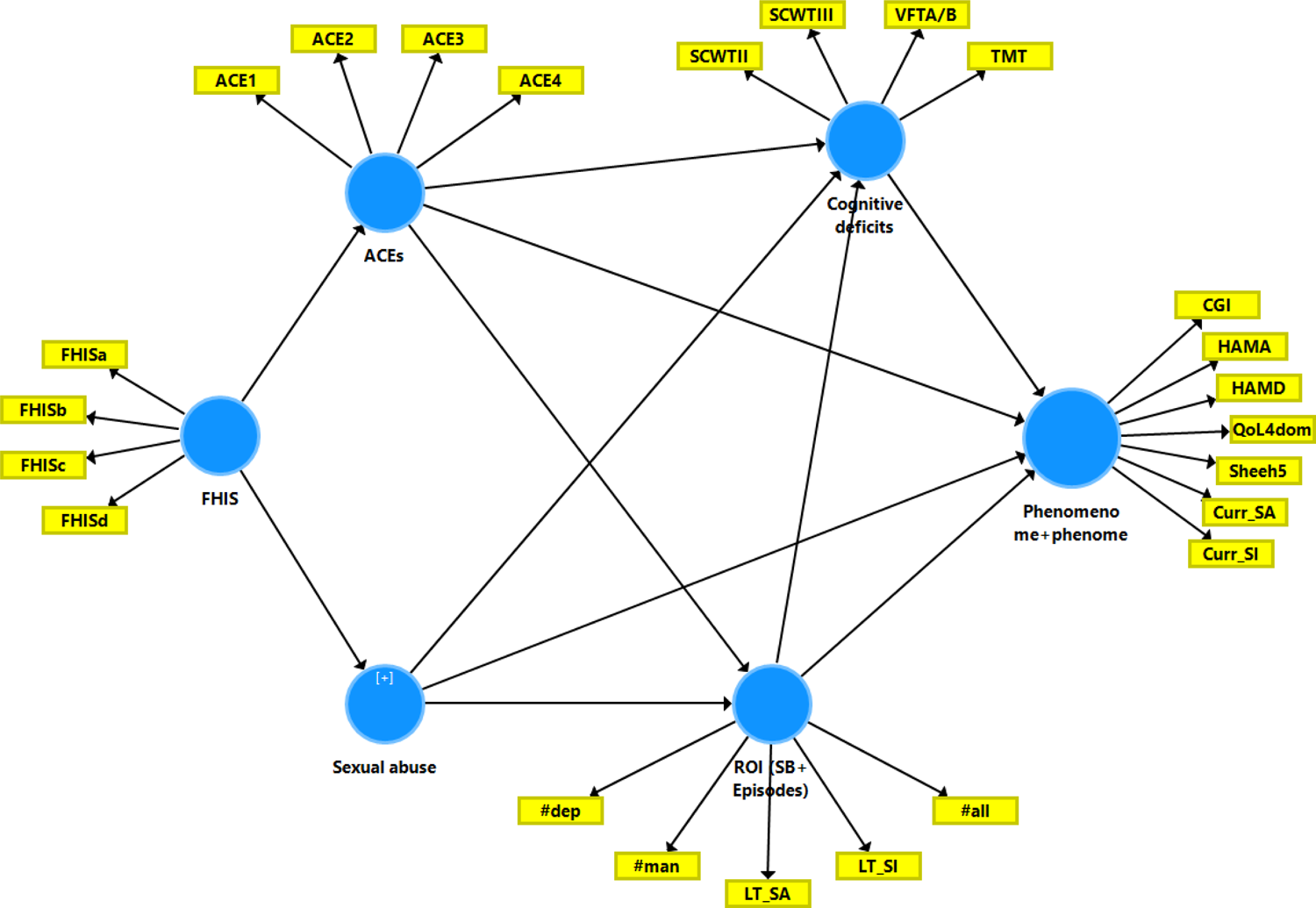
Theoretical framework showing the causal links between the building blocks of mood disorders. Output (dependent) variable is the phenome-phenomenome score comprising the Hamilton Depression and Anxiety Rating Scale (HAMD/HAMA) scores; four quality of life domains (QoL4dom); five disability Sheehan (Sheeh5) scores; current suicidal ideation and attempts (Curr_SI and Curr_SA, respectively); and the Clinical Global impression (CGI) score. Explanatory variables are recurrence of illness (ROI), namely a combination of lifetime suicidal behaviors (SB) and depressive (#dep) and (hypo)mania (#mania) episodes; adverse childhood experiences (ACE1-4) and sexual abuse; and a family history (FHIS) of mood disorders, suicidal attempts and different substance abuse disorders; and cognitive deficit tests including trail making test (TMT) Stroop test (SCWT) and Verbal fluency test (VFT).

Complete PLS path analysis was performed only when the inner and outer models matched the following predetermined quality criteria: a) the overall model fit, namely the standardized root square residual (SRMR) value is <0.08, b) the latent vectors of the outer models demonstrate high convergence and construct validity as indicated by Cronbach’s alpha > 0.7, composite reliability > 0.8, rho A > 0.8 and average variance extracted (AVE) > 0.5, and loadings >0.6 on the extracted latent vectors at p < 0.0001, c) blindfolding demonstrates that the construct’s cross-validated redundancies (CCVR) or communalities (CCVC) are sufficient, d) Confirmatory Tetrad Analysis (CTA) confirms that the latent vectors are not misspecified as reflective models, and e) the model’s prediction performance as tested by PLS Predict is satisfactory. Prediction-Oriented Segmentation (POS)-PLS is performed to establish measurement invariance and Multi-Group Analysis (MGA)-PLS to decipher whether the constructs stem from a single population, thereby excluding population heterogeneity.

In this paper we will describe AVE and Cronbach alpha as well as CCVR/CCVC but the complete model quality data can be obtained from the corresponding author (as html files). In all cases, CFA showed that the models were not mis-specified as reflective models. If all the above-mentioned model quality data met the predetermined criteria, we conducted a complete PLS path analysis with 5,000 bootstrap samples, produce the path coefficients (with exact p-values), and additionally compute the specific indirect and total indirect (that is, mediated) effects in addition to the total effects.

## Results

### Construction of suicidal behavior latent vectors

Based on the causal framework presented in Figure 1 we first delineated the effects of FHIS and ACE data on Y5 SA and current SI data and determined the mediated paths by LT SI and LT SA data, see **Electronic Supplementary File (ESF)**, Figure 1. We extracted a common factor from emotional/physical abuse and neglect (dubbed: ACEphem, AVE=0.721, Cronbach alpha=0.872, all loadings > 0.772, CCVR=0.060). Sexual abuse was entered as a single indicator. A common factor could be extracted from FHIS5 (MDD), FHIS6 (BD) and FHIS7 (SA) (labeled FHISmood) (AVE=0.645, Cronbach alpha=0.811, all loadings > 0.665, CCVC=0.436) and from FHIS2 (TUD), FHIS3 (alcohol dependence) and FHIS4 (SUD) (dubbed FHISabu; AVE=0.618, Cronbach alpha=0.777, all loadings > 0.654, CCVC=0.400).

We extracted one latent vector from 10 LT SI items (AVE=0.741, Cronbach alpha=0.961, all loadings > 0.803, CCVR=0.160) and from 5 LT SA items (AVE=0.566, Cronbach alpha=0.809, all loadings > 0.657, CCVR=0.226), and as such these latent vectors reflect LT SI and LT SA, respectively. Moreover, we extracted latent vectors from five C-SSRS items of 5Y SA (AVE=0.628, Cronbach alpha=0.851, all loadings > 0.653, CCVR=0.096).

ESF, Figure 1 shows the PLS model with an SRMR=0.054, indicating adequate model fit. We found that 24.3% of the variance in current SI was predicted by LT SI and ACEphem. We found that 18.5% of the variance in 5Y ACE was predicted by LT SI and FHISabu. Up to 22.1% of the variance in LT SI was predicted by ACEphem, sexual abuse, whilst LT SI explained 42.7% of the variance in LT SA. There were specific indirect effects of FHISmood on current SI mediated by ACEphem (t=2.26, p=0.024) and the path from ACEphem to LT SI (t=2.59, p=0.010) yielding a significant total effect (t=0.02, p=0.003). There were also significant specific indirect effects of ACEphem (t=3.64, p<0.001) (but not sexual abuse) on current SI that were mediated by LT SI. As such, FHISabu predicted LT SA, whilst FHISmood predicted LT/current SI.

In order to construct a RADAR score reflecting overall suicidal behaviors (SB) we have extracted a latent vector from LT SI, LT SA, current SI, and 5Y SA. We found that 44.8% of the variance in this overall SB score was predicted (F=31.63, df=4/156, p<0.001) by number of depressive episodes (β=0.469, t=7.27, p<0.001), ACEphem (β=0.172, t=2.44, p=0.016), FHISabu (β=0.142, t=2.26, p=0.025) and sexual abuse (β=0.139, t=2.08, p=0.039).

### Construction of ROI latent vectors

**ESF Figure 2** builds on the results of ESF, figure 1 and includes a new ROI1 index, constructed using LT number of depression, (hypo)manic and total episodes (AVE=0.828, Cronbach alpha=0.897, all loadings > 0.835, CCVR=0.155). We found that 19.7% of the variance in this ROI1 index was predicted by ACEphem and FHISmood and that 32.6% of the variance in LT SI was predicted by ACEphem and ROI1. The latter had also direct effects on LT SA and together with LT SI predicted 24.3% of the variance in current SI, indicating that LT SI mediated the effects of ROI on current SI (t=3.48, p=0.001). Also, the model shown in ESF, Figure 2 displays accurate model fit with SRMR=0.052.

**Figure 2.**
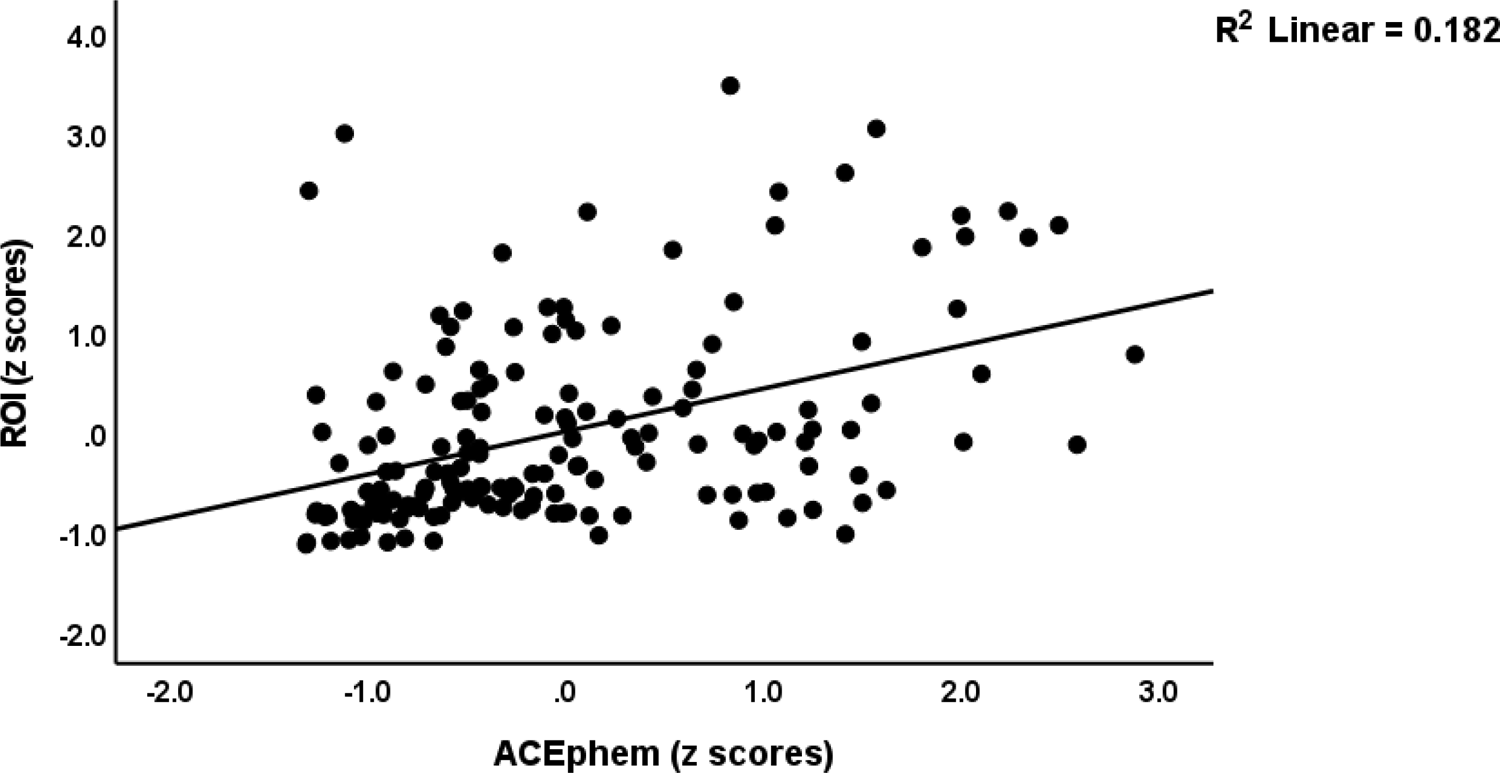
Partial regression of the recurrence of illness index on adverse childhood experiences (physical and emotional neglect and abuse)

**ESF, Figure 3** builds on the two previous models and aims to construct the second ROI2 index (number of depression and mania episodes and LT SI and SA latent vectors), a cognitive vector (combining SCWT, TMT and VFT scores) and a latent vector that combines current clinical data (HAMD, HAMA and CGI) with phenomenome (WHO-Qol and Sheehan disability) data. This figure shows that we extracted one validated ROI2 factor comprising number of depression an manic episodes and the LT SI and SA latent variables (AVE=0.630, Cronbach alpha=0.849, all loadings > 0.688, CCVR=0.155). On the other hand, no validated latent factor could be extracted from the cognitive test scores. Consequently, we computed a z-unit-based composite score as z value of the first factor extracted from the STROOPI, II and III scores (explaining 75.5% of the variance, all loadings >0.830) – z (z VFTA+z VFTB) (dubbed CogFLEX). The latter was entered as a single indicator in the PLS analysis. We also extracted one latent vector from the clinical and self-rating scores, dubbed phenome1 (AVE=0.689, Cronbach alpha=0.888, all loadings > 0.785, CVR=0.269).

**Figure 3.**
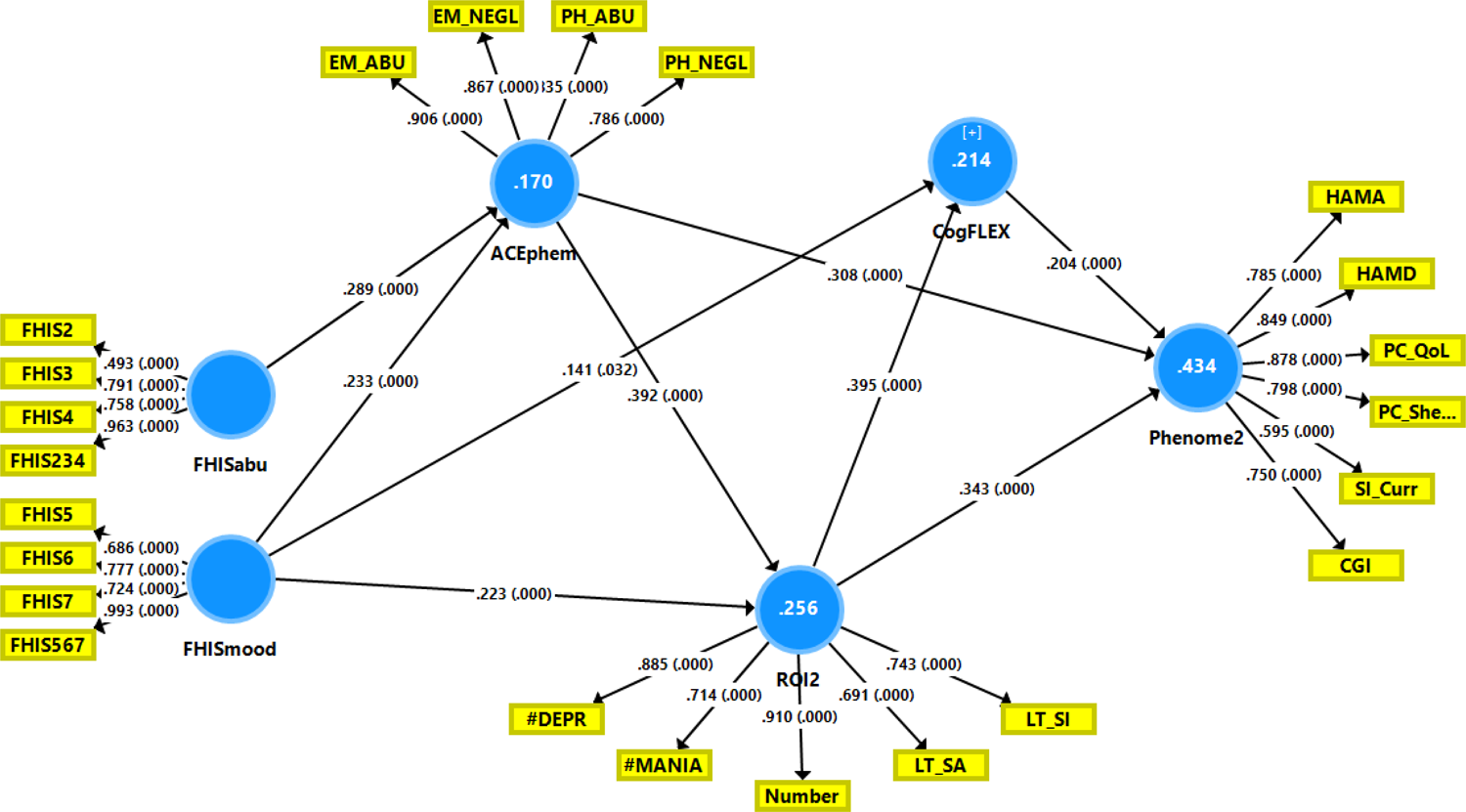
Final Partial least squares model of mood disorders. This model shows the causal pathways from a family history of mood (FHISmood) and substance use (FHISabu) disorders, to adverse childhood experiences (ACEphem) physical and emotional abuse (PH_ABU and EM_ABU) and physical and emotional neglect (PH_NEGL and EM_NEGL), to recurrence of illness (ROI), namely a combination of lifetime number of depressive (#dep) and (hypo)mania (#mania) episodes and suicidal ideation (LT_SI) and attempts (LT_SA); to cognitive deficits in verbal fluency and executive functions (CogFLEX). The final outcome variable is an integrated index of the phenome and phenomenome, comprising the Hamilton Depression and Anxiety Rating Scale (HAMD/HAMA) scores; four quality of life domains (QoL4dom); five disability Sheehan (Sheeh5) domain scores; current suicidal ideation (Curr_SI); and the Clinical Global impression (CGI) score. Shown are the significant paths, path coefficients (with p values) and the loading (with p values) of the outer models. Figures in the blue circles denote the explained variances.

With an SRMR of 0.052 the model fit shown in ESF Figure 3 was more than adequate. This model shows that 42.2% of the variance in the phenome1 score was predicted by the combined effects of CogFLEX, ROI2, ACEphem, FHISmood and FHISabu, whilst 27.0% of the variance in current SI was explained by ROI2 and ACEphem. As such, ROI2 had significant direct effects on phenome1 and current SI (see figure 4) and indirect effects on phenome1 (t=2.64, p=0.004) that were mediated via CogFLEX. Moreover, ROI2 mediated the effects of FHISmood and ACEphem on phenome1 (t=2.45, p=0.007) and CogFLEX (t=2.88, p=0.002). **Figure 2** shows the partial regression of the ROI2 score on ACEphem (after adjusting for age and sex).

**Figure 4.**
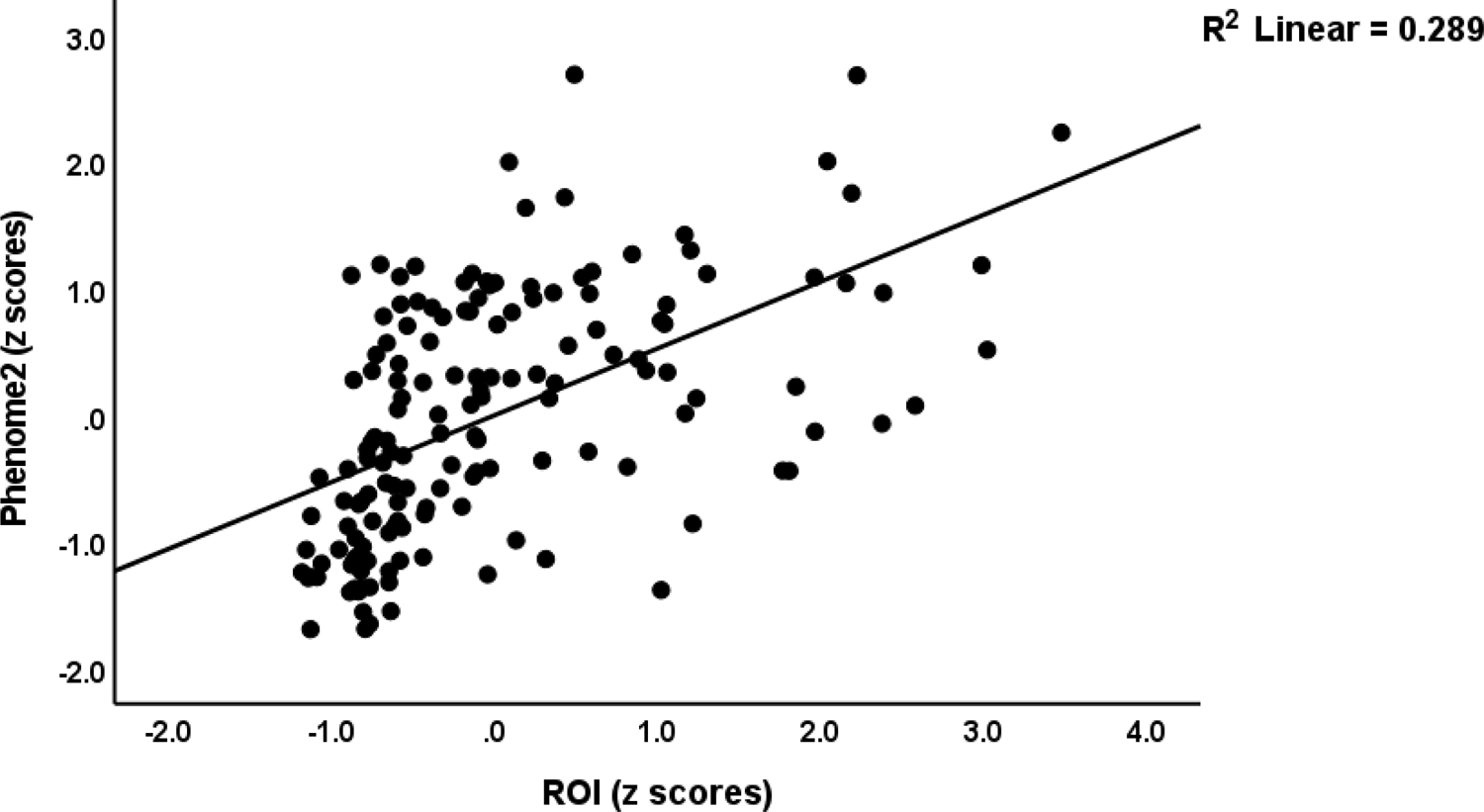
Partial regression of the phenome of mood disorders on recurrence of illness (ROI)

### Final PLS model

The next question was whether the latent vector extracted from the 9 current SI indicators could be incorporated into the phenome1 construct. **Figure 3** shows that we were able to extract one factor from the 5 clinical phenome1 data and the latent variable extracted from the current C-SSRS items (AVE=0.611, Cronbach alpha=0.869, all loadings > 0.595, CCVR=0.254). With an SRMR of 0.057, the model fit was adequate. This final model shows that 43.4% of the phenome2 (final output variable) is predicted by the regression on ROI2, ACEphem and CogFLEX, and that 21.4% of the variance in CogFLEX was explained by ROI2 and FHISmood. Moreover, 25.6% of the variance in ROI2 was predicted by ACEphem and FHISmood, whereas 17.0% of the variance in ACEphem was explained by the combined effects of FHISmood and FMISabu. All the possible indirect (mediated) paths from the input variables (FHIS and ACE data) and ROI2 to phenome2 were significant, except for the path from FHIS to phenome mediated by CogFLEX (NS at p=0.057). Total effects showed that phenome2 was highly significantly predicted by FHISmood (5.11, p<0.001), FHISabu (3.47, p<0.001), ACEphem (t=8.57, p<0.001), ROI2 (6.53, p<0.001) and CogFLEX (t=3.82, p<0.001). **Figure 4** shows the partial regression of the phenome2 score on the ROI2 score, and **Figure 5** shows the partial regression of the phenome2 score on ACEphem (both after adjusting for age and sex). Post-hoc minimum sample size estimation shows that to obtain a power of 0.9 at p=0.05, less than 173 individuals should be included for all paths.

**Figure 5.**
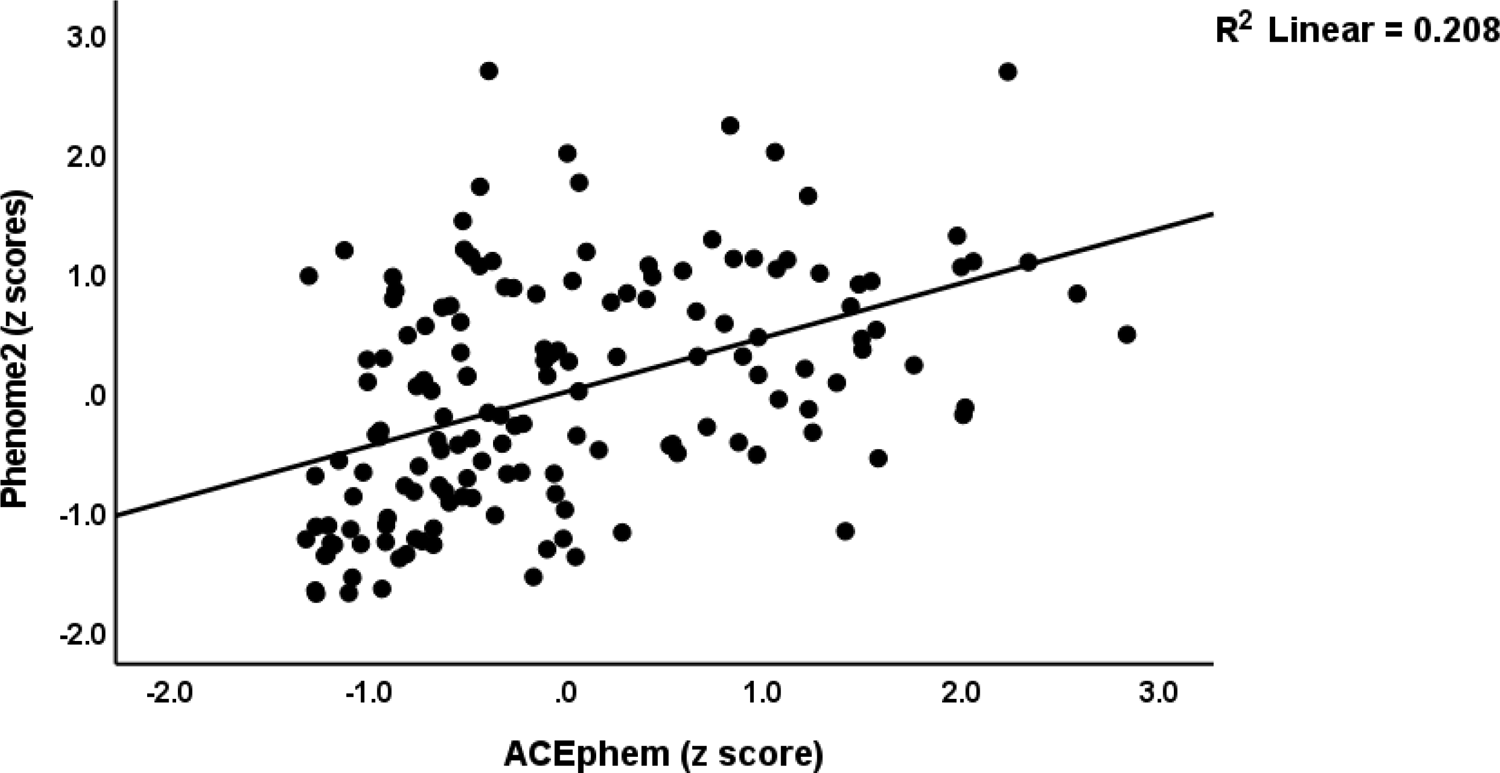
Partial regression of the phenome of mood disorders on adverse childhood experiences (physical and emotional neglect and abuse)

### PLS Multigroup analysis (MGA) and Prediction Oriented Segmentation (POS)

To exclude significant group heterogeneity or measurement invariance we performed MGA-PLS and PON-PLS on the final model (Figure 3). The major aims were to examine whether there are differences in the path coefficients between BD and MDD and whether the BD group was detected using POS. MGA comparing BD and MDD showed that there were no significant path-coefficient differences between both study groups. For example, the paths from ROI2 to phenome2 were equally (p=0.308) significant in MDD (t=6.62, p<0.001) and BD (t=4.06, p<0.001); the paths from ACEphem to phenome2 were equally significant (p=0.332) between MDD (t=3.71, p<0.001) and BD (t=3.92, p<0.001); and also the paths from ACEphem to ROI2 were equally significant (p=0.134) between MDD (t=3.71, p<0.001) and BD (t=3.53, p<0.001). We also ran PLS-MGA using sex, TUD, MetS, and any FHIS as grouping variables, but again no significant path-coefficient differences could be detected. PLS-POS with phenome2 as target showed no measurement variance when considering the ROI2 and phenome 2 associations.

### Construction of a ROI-phenome score

We extracted a validated factor from ROI2, and latent vectors extracted from the three clinical data (HAMD, HAMA, CGI), and 4 WHO-QoL and 5 Sheehan domains (AVE=0.659, Cronbach alpha=0.840, all loadings>0.750, CCVR=0.400). Using a visual binning method, this ROI-phenome score was binned into 3 groups using cut-off values of −0.61 and +0.23. Those with a score <-0.61 were the normal controls, the group with scores between <-0.61 and >0.23 was called simple dysmood disorder (SDMD), and the group with scores > +0.23 was labeled major dysmood disorder (MDMD).

ESF Table 1 shows that there were no significant differences in age, sex, BMI, TUD, and MetS between the HC, SMDM and MDMD groups while education was marginally lower in patients than controls. **Figure 6** shows the bar graph with the key features of both depression subgroups as compared with controls. Sexual abuse was significantly higher in MDMD than in controls (p<0.001) and SDMD (p=0.009). FHIS567 (all at p<0.001), ACEphem (all at p<0.007), ROI2 (all p<0.001) and phenome2 (all p<0.001) were significantly different between the three groups and increased from healthy controls → SDMD → MDMD.

**Figure 6.**
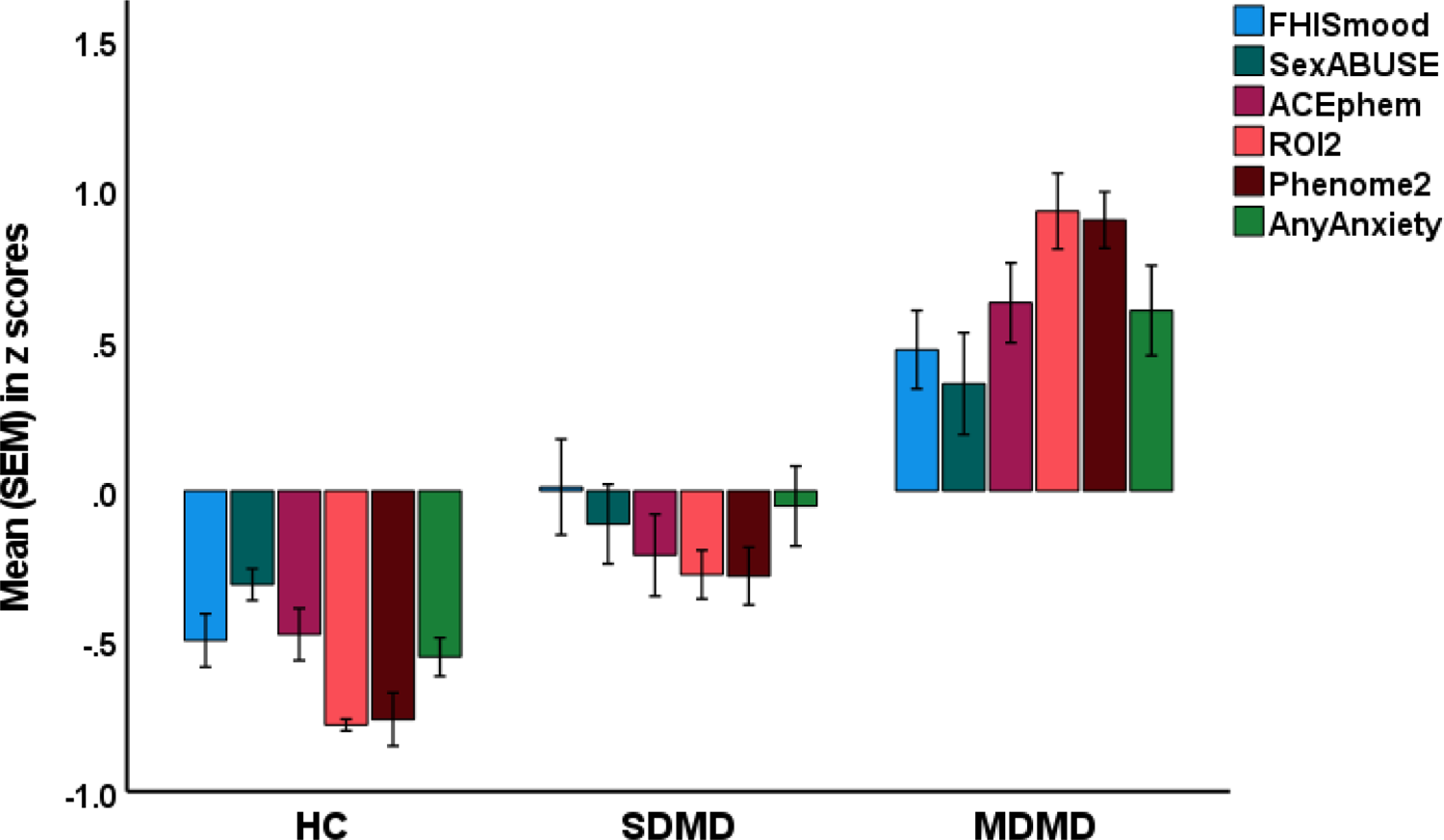
Clustered bar graph showing the RADAR score differences between healthy controls (HC) and patients with mood disorders divided into those with major (MDMD) and simple (SDMD) dysmood disorder. FHISmood: a family history of mood disorders and suicide; ACEphem: adverse childhood experiences (neglect and abuse); ROI2: recurrence of illness index; phenome2: an index of phenome and phenomenome; any anxiety: number of comorbid anxiety disorders.

**Figure 7** shows the RADAR plot displaying the mean RADAR scores of SMDM and MDMD patients. This graph shows the relative position of the mean values obtained in MDMD/SDMD (expressed in standard deviations multiplied by 10) versus the common center point, which was defined as the mean value of all feature scores of healthy controls set to 0. There are 12 axes, and each axis represents one feature score. The graph also connects the axes, so separating the graph into grids that indicate the difference between the feature scores of patients and controls. The number of depression and manic episodes, LT SI, LT SA, 5Y SA score, and current SI were significantly higher in MDMD than in controls and SDMD. The number of comorbid anxiety disorders was significantly higher in MDMD than in SDMD (p<0.013) and controls (p<0.001).

**Figure 7.**
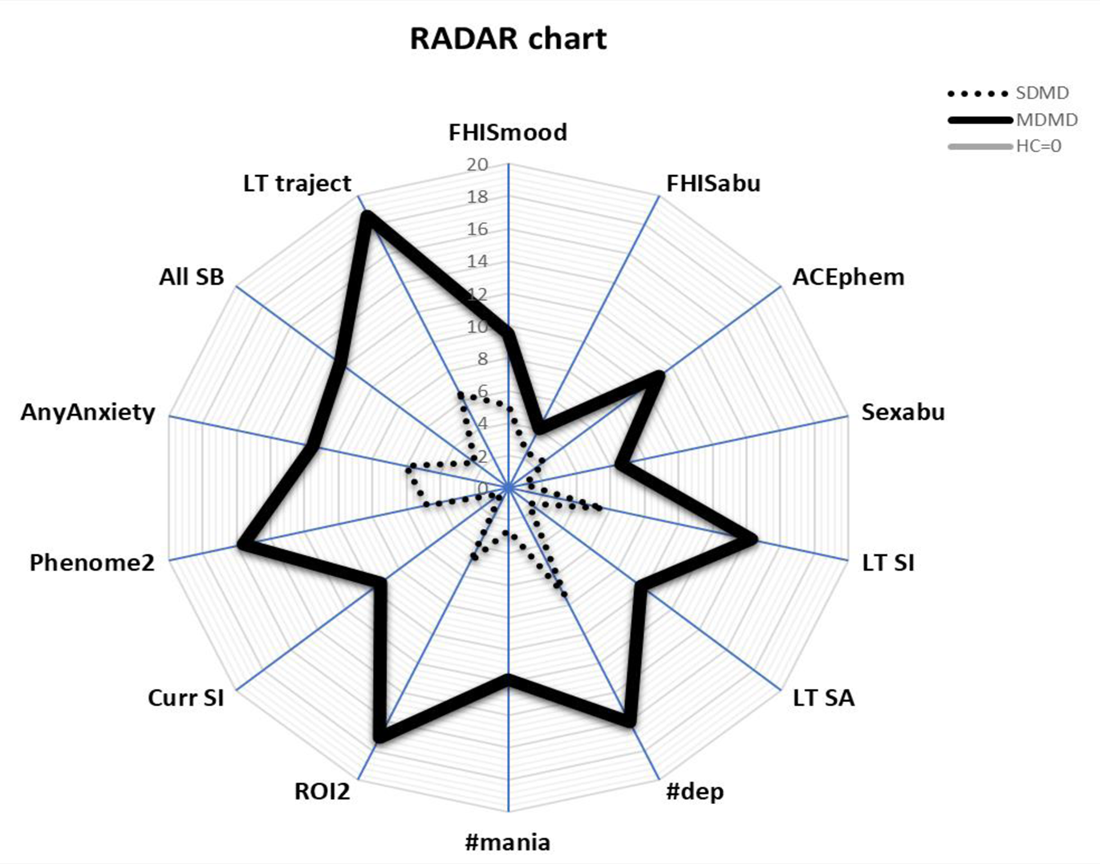
RADAR graph displaying the Research and Diagnostic Algorithmic Rules (RADAR) scores. HC: healthy controls (set at 0); SDMS/MDMD: simple and major dysmood disorder. FHISmood/FHISabu: family history of mood/suicide and substance use disorders, respectively; ACEphem: adverse childhood experiences (physical and emotional neglect and abuse); Sexabu: adverse childhood experiences, sexual abuse; LT SI/SA: lifetime suicidal ideation and attempts, respectively; #dep: number of depressive episodes; #mania: number of lifetime (hypo)manias; ROI2: recurrence of illness index; Curr SI: current suicidal ideation; Phenome2: the phenome-phenomenome of mood disorders; AnyAnxiety: number of comorbid anxiety disorders; All SB: an index of all lifetime and current suicidal behaviors (SI and SA) combined; LT Traject: lifetime trajectory score. This graph shows the relative position of the subject versus the common center point. The latter was defined as the mean value of all features of the healthy controls set to 0. All feature scores are displayed in standard deviations (multiplied by 10) on 12 axes whereby each axis represents one feature score. These graphs also connect the axes, so separating the graph into grids that indicate the difference between the feature scores of patient groups and controls.

Consequently, we have delineated the best prediction of the ROI2-phenome2 score using the most accurate selection of predictors. Using 8 predictors, we found that 94.9% of the variance could be predicted (F=332.20, df=8/144, p<0.001) by total number of episodes (unstandardized B=0.052), HAMD (B=0.043), CGI (B=0.150), C-current C-SSRS1 (B=0.209, wish to be death), current C-SSRS4 (B=0.287, suicidal intent), LT C-SSRS2 (B=0.214, suicidal thoughts), LT C-SSRS5 (B=0.253, with planning) and 5Y C-SSRS12 (B=0.033, number suicidal attempts last 5 years), whist the constant is −1.383. Consequently, we performed ROC analysis on the standardized predicted values to delineate the best threshold value to separate MDMD from SMDM. The coordinate tables of the ROC curve (area under the ROC curve=0.991 ±0.005) showed that the best cut-off value of the unstandardized predicted value was 0.2635244 (sensitivity=93.3% and specificity=100%). This indicates that the computed regression model based on a reduced number of easily measurable variables performs well in discriminating MDMD from SDMD and that this algorithm may be used to make the diagnosis of MDMD.

### Towards a mood disorder lifetime trajectory score

We also extracted a replicable and validated latent vector from ROI2, phenomenome2, ACEphem and any anxiety disorder (AVE=0.578, Cronbach alpha=0.756, all loadings>0.726, CCVR=0.121). Multiple regression analysis showed that 21.2% of the variance in this traject score (F=13.11, df=3/146, p<0.001) was predicted by the regression on female sex (β=0.177, t=2.36, p=0.019) FHISmood ((β=0.327, t=4.29, p<0.001) and FHISabu (β=0.159, t=2.11, p=0.037). A visual binning method divided the sample into four study groups, namely controls, and phase 1, phase 2 and phase 3. ESF Figure 4 (bar graph) summarizes the key characteristics of the three phases. Phase 3 patients showed increased FHISmood, sexual abuse, ACEphem ROI2, and lifetime traject scores as compared with phase 1 + phase 2 patients. Moreover, the prevalence of panic disorder (χ^2^=16.47, df=1, p<0.001), OCD (χ^2^=5.35, df=1, p=0.021), PTSD (χ^2^=6.66, df=1, p=0.010), social phobia (χ^2^=7.62, df=1, p=0.006), and simple phobia (χ^2^=6.39, df=1, p=0.01), but not GAD (χ^2^=1.47, df=1, p=0.225) was higher in phase 3 than in phase 1 + phase 2 patients. The prevalence of any anxiety disorder was significantly higher (χ^2^=29.82, df=1, p<0.001) in phase 3 than in phase 1 + phase 2 patients.

### Clinical useful algorithms

ESF, Tables 2-6 show the results of 5 (bootstrapped, n=1000) multiple regression analysis with ROI (ESF 2, Table 2), LT SB (ESF, Table 3), current SI (ESF, Table 4), phenome 2 (ESF, Table 5) and the lifetime trajectory score (ESF, Table 6) as dependent variables (always entered as z scores) and their single indicators (not the extracted factors because they are not clinically useful) as explanatory variables. ROI2 was best predicted by total number of episodes, C-CSSR-6 LT and C-CSSR-12 LT (F=1984.10, df=3/161, p<0.001, effect size=0.974). The LT SB score was best predicted by C-SSRS LT items 2, 4, 5, 6, 7 and 12 (F=214.59, df=6/159, p<0.001, effect size=0.890). The current suicidal ideation score was best predicted by the C-SSRS-current items 1, 3 and 6 (F=1882.03, df=3/163, p<0.001, effect size=0.9714). The phenome2 score was best predicted by C-SSRS-current items 1 and 4, HAMD, HAMA and CGI scores (F=248.47, df=5/149, p<0.001, effect size=0.893). The lifetime trajectory score was best predicted by number of depressive episodes, emotional abuse, emotional neglect, HAMD, Sheehan 1 (work/school) score and any anxiety disorder (yes/no) (F=397.72, df=6/144, p<0.001, effect size=0.943). ESF Table 7 shows the Q50 and Q75 values, demarcating the more severe and very severe cases.

### Effects of the drug state

ESF, Table 8 demonstrates the point-biserial correlations between the drug state variables and FHISmood, ACEphem, ROI2, and phenome2, with most variables exhibiting significant intercorrelations. Multiple regression analysis reveals that the use of antidepressants (β=0.253, t=3.36, p=0.001), antipsychotics (β=0.170, t=2.11, p=0.037), and mood stabilizers (β=0.266, t=2.80, p=0.006) predicted 19.5% of the variation (F=11.60, df=3/144, p=0.001) in the phenome2 scores. Nonetheless, the impact of the drug state variables was no longer significant after including ACEphem (β=0.275, t=3.71, p<0.001) and ROI2 (β=0.288, t=3.50, p<0.001) in the analysis.

## Discussion

### RADAR scores and plots

The primary outcome of this work is the development of Research and Diagnostic Algorithmic Rule (RADAR) scores that reflect the key aspects of mood disorders, including ACE, ROI, phenome, and suicidal behaviors. In addition, we explained how to plot these scores in RADAR graphs (or star or spider graphs), which display all features of the disease (multivariate data translated into algorithms) in a two-dimensional plot. These graphs permit a) the comparison of the unique characteristics of two or more patients or phenotypes, e.g. MDMD versus SDMD (**Figure 7**), and b) the identification of the major characteristics of a particular patient.

In addition, the current study proposes a diagnostic algorithm for MDMD, a new data-driven mood disorder phenotype (Maes et al., 2021a; Simeonova et al., 2021). This phenotype is based on a comprehensive evaluation of the phenome, which includes depression, anxiety, and global clinical ratings, as well as disabilities and HR-QoL, and ROI scores, including recurrence of episodes and suicidal tendencies. This diagnostic group is defined not only by more severe symptoms and disabilities and a worse HR-QoL but also by a higher ROI and is externally validated by higher ACE ratings and familial histories of mood disorders and SUDs. Previously, we have shown that this phenotype is substantially related with enhanced cytokine and growth factor network responsiveness, increased nitro-oxidative stress, genetic variants in the PON1 Q192R gene, autoimmunity and hypernitrosylation (Maes et al., 2018; 2019; 2021a; 2021b; 2022c; Simeonova et al., 2021), whereas SDMD is not associated with such pathways. Our new RADAR approach, which synthesizes multivariate diagnostic data into useful algorithms that score all features of mood disorders, contrasts with the gold standard psychiatric approach to diagnose mood disorders with DSM/ICD criteria and rate the severity of illness with a rating scale (e.g., HAMD). The DSM/ICD diagnoses have a major flaw in that patients with SDMD (not associated with biomarkers) and MDMD (strongly associated with biomarkers) are lumped together with patients with distress responses, grief, and demoralization, resulting in extreme heterogeneity and thus incorrect models of mood disorders. As discussed in the Introduction, mood disorder diagnoses based on DSM/ICD case definitions are invalid, and there is often low agreement among psychiatrists, with certain subtypes being under-or overvalued (Maes, 2022; Maes and Stoyanov, 2022). As a result, it is preferable to employ our quantitative scores, including the phenome and ROI, and MDMD (a homogenous class that is strongly associated with biomarkers) as dependent variables in statistical analyses rather than an inaccurate DSM/ICD model (most often used as an explanatory variable). In addition, while our RADAR scores are derived using bottom-up machine learning methods, the DSM/ICD case definitions are generated using a top-down method based on a consensus among so-called professionals (Maes, 2022; Maes and Stoyanov, 2022). As described previously, our RADAR model is falsifiable, confirmable, truth-value determinable, and also progressive and changeable (Popper, 1962; Antiseri, 1991), whereas the dogmatic structure of DSM/ICD prevents diagnostic concept falsification, therefore preventing deductive and inductive learning (Maes, 2022; Maes and Stoyanov, 2022).

The DSM/ICD criteria’s consequence is even more bizarre: whereas our RADAR scores defining MDMD should be utilized as a dependent (outcome) factor in statistical studies, the majority of researchers employ the gold standard DSM/ICD as explanatory (input) variables. As a result, the vast majority of research uses an inaccurate and controversial higher-order construct together with incorrect model assumptions (input and output variables are reversed), which are then analyzed with improper statistical tests (Maes, 2022). As previously stated, most psychiatric rating scales are inaccurate constructs (namely not based on a common factor and not unidimensional) that are founded on folk psychology-like narratives (Maes, 2022; Maes and Stoyanov, 2022).

Even more repellent is the American Psychiatric Association’s and folk psychologists’ claim (APA, 2013; Zachar and Kendler, 2017) that decades of biological research failed to identify solid biomarkers of mood disorders, despite the fact that the most important requirement for statistical tests and machine learning was never met due to their own failures not to provide a correct model (Maes, 2022). Statistical analyses are inaccurate and should not be performed without a correct (that is, reproducible and cross-validated) disease model. Nonetheless, the Western status quo in psychiatry is far from admitting the inadequacies in their methodology. When submitting a research manuscript, for example, the Lancet Psychiatry recommends establishing one primary measurement (Boyce et al., 2021). This says it all: the outcome variable should be a correct model, not an erroneous or folk-psychology-derived measurement. Even worse, the Lancet calls for a “primary” measurement, as if one must choose between two or more incorrect measurements. The current paper’s message, however, is clear: rather than a single selected measurement, a composite or latent vector generated from several phenome and phenomenome data should be employed as the final output variable.

Consequently, in pan-omics biomarker research, our quantitative RADAR scores should be used rather than the invalid DSM/ICD diagnoses or a folk-psychology-generated rating scale score. Moreover, future biomarker research should utilize our quantitative RADAR scores and the MDMD criteria to develop novel pathway phenotypes (RADAR combined with biomarker data) and endophenotype classes (biomarkers combined with MDMD criteria) to predict prognosis, treatment responsiveness, remission, and treatment resistance (Maes, 2022). Moreover, while MDMD is clearly distinguished from SDMD, our analysis of lifetime trajectory revealed that the intermediate class between MDMD and controls can be further subdivided into two patient classes (phase 1 and phase 2). Importantly, future research should establish new diagnostic criteria to distinguish normal distress reactions (sadness and demoralization) from phase 1 patients with a first episode who will eventually develop MDMD. This approach will necessitate the use of genetic markers as well as gene X environmental (ACE) interactions.

### RADAR scores assessing suicidal behaviors (SB)

Suicidal behavior scores are important components of the phenome of mood disorders, ROI and MDMD in our study, and we also generated new data-driven scores of lifetime and current suicidal ideation and attempts as well as overall suicidal behaviors. The latter was strongly predicted (44.8% of its variance) by number of depressive episodes, ACEphem and sexual abuse, and a family history of SUDs. In addition, our PLS analysis revealed that 27% of current SI was predicted by the combined effects of ROI and ACEphem, and that ACEphem completely mediated the effects of a family history of mood disorders on current SI. Moreover, our models demonstrated that current suicidal ideation share the same common core with severity ratings of sadness and anxiety, disabilities and HR-QoL. In accordance with a recent study (Maes et al., 2022) demonstrating that, in mood disorders, a replicable and validated latent factor can be extracted from recent suicidal behaviors, psychotic and melancholic features, and the HAMDD and STAI scores, the current findings indicate that a similar pathophysiology underlies these phenome characteristics. Recent meta-analyses demonstrated that active immune-inflammatory and nitro-oxidative pathways significantly underlie lifetime and recent suicidal behaviors (Vasupanrajit et al., 2021; 2022). Recent research indicates that elevated M1 macrophage, T helper 1, and T cell growth factor responsiveness is substantially linked with lifetime and current suicidal behaviors (Maes et al., 2022b).

Nevertheless, the present study also established differences between the types of suicidal behaviors. Thus, ACE and a family history of mood disorders predicted lifetime and current suicidal ideation, whereas lifetime and more recent suicidal attempts are predicted by a family history of SUDs. Likewise, previous studies demonstrated that alcohol misuse and TUD are related to suicidal attempts and that these correlations remained substantial even after depression was accounted for (Wu et al., 2004). In contrast, there was no longer a correlation between suicidal ideation and SUDs when depression was controlled for (Wu et al., 2004). Previous meta-analyses have revealed discrepancies in the relationships (effect sizes) between immune-oxidative pathways and current vs. lifetime suicidal behaviors, as well as between suicidal ideation vs. attempts (Vasupanrajit et al., 2021; 2022). However, as the latter systematic review showed, earlier research has frequently grouped together attempts and ideation, and a lifetime history with current behaviors. Nevertheless, our data indicate that it is crucial to distinguish between these distinct concepts because they contribute differentially to ROI (lifetime assessments), the overall severity of the illness (current behaviors), and the lifetime trajectory score (lifetime and current), and thus to the extremely high rate of suicidal ideation among phase 3 patients.

### ROI scores

In our research, the ROI index was developed as a reflective latent factor with adequate replicability and reliability validity that was generated from the number of depressive and (hypo)manic episodes, as well as lifetime suicidal ideation and attempts; as such, it is a critical component of our mood disorders model. Again, this shows that recurrence of episodes and suicidal behaviors are manifestations of a common single trait, namely ROI, and they share a common pathophysiology. In the present study, we found that ACEphem and a family history of mood disorders are significant predictors of ROI. Previous research has shown that ROI is strongly predicted by gene (paraoxonase 1 or PON1 Q192R gene) X ACE interactions and the resulting deficits in PON1-high density lipoprotein (HDL) cholesterol complex (Maes et al., 2018; 2019), as well as with an increased immune and growth factor responsiveness in unipolar depression (Maes et al., 2022b), and dysfunctional T regulatory activities in bipolar disorder (Maes et al., 2021b). Interestingly, the frequency of seizures (kindling) in temporal lobe epilepsy is related with oxidative pathways and decreased antioxidant defenses, particularly PON1 activity and sulfhydryl groups (Maes et al., 2022a). Previous research revealed that oxidative and inflammatory indicators, as well as specific pro-inflammatory cytokines, predicted the recurrence of episodes, hospitalizations, and suicide attempts (Maes et al., 2001; Celik et al., 2010; Maes et al., 2012; Grande et al., 2014; Hope et al., 2013; Siwek et al., 2017; Sowa-Kucma et al., 2018). These links were thought to be the result of sensitization of neuro-immune and oxidative stress pathways (Maes et al., 2001; 2012).

Our study found that ROI predicts current suicidal ideation, cognitive deficits, and the phenome of mood disorders, suggesting that ROI is the major driver of the severity of the phenome and that it mediates the effects of genetic, environmental, and biological factors on the latter. Future research should therefore examine associations between genetic and other biomarkers and a patient’s quantitative ROI score, rather than focusing on correlations between (genetic) biomarkers and an incorrect DSM/ICD diagnosis. These correlations will reveal which genetic factors, alone or in combination with ACEs, are responsible for ROI, the phenome, and ultimately increased suicide behavior.

In contrast to our comprehensive and quantitative ROI score, the DSM and ICD only permit the diagnosis of single and recurrent depression, despite the fact that it is obvious from our analysis that a high ROI score, and not recurrence per se, is the primary driver of severe mood disorders. Recent initiatives, such as the Timeline Course Graphing Scale for the DSM-5 Mood Disorders (TCGS) (Mccullough et al., 2016) and the NIMH Life Chart Method (LCM-p) (Denicoff et al., 2002), intend to depict the course of illness but do not seek to compute scores that reflect ROI or the severity of ROI as a driver of the phenome. Some authors (Berk et al., 2011; Kapczinski et al., 2009; Ferensztajn et al., 2014) have proposed models for staging and offered theoretical staging models that, unlike ours, were not based on patient data and were viewed as ordinal data rather than scale scores.

A new systematic review discusses unipolar depression staging models and finds that the majority of studies focused on treatment resistance (Cosci and Fava, 2022). One model, for example, suggests a stage paradigm based on failure to respond to adequate trials with psychotherapy and antidepressants. The question remains, however, what constitutes an adequate therapeutic trial with psychotherapy and antidepressants. Thus, the model developed here demonstrates that ROI (and the pathophysiology behind ROI) drives suicidal behaviors and the phenome and thus treatment resistance, both in the acute phase of depression (as indicated by the severity of the phenome) and over time (as indicated by ROI). One can wonder how psychotherapy could affect the major neuro-immune and neuro-oxidative pathophysiology, which includes genetic polymorphisms (PON1 Q192R genotype) sensitization of the immune network, T regulatory dysfunctions, autoimmunity and hypernitrosylation (Maes et al., 2018; 2019; 2022a; 2022b). As discussed in our limitations section, antidepressants have no effect on ROI, and furthermore, antidepressants have extremely detrimental effects on the immune system, causing increased vulnerability to injuries and stress (Maes et al., TBS). Another model mentioned by Cosci and Fava (2022) divides unipolar depression into five stages: the first is a prodomal stage; the second is a first episode; the third is a residual phase (stage 3); the fourth is recurrent or double depression; and the fifth is chronic MDD. Nonetheless, this theoretical model suggests some top-down categories that lack information on the key features of mood disorders, particularly ROI and the phenome. Moreover, none of the models addressed in the study are bottom-up models built and cross-validated with patient data and machine learning, and the only data-driven models of mood disorders that contain biomarker data (Maes et al., 2018; 2019; 2021b) are not even mentioned. As a result, the only precision medicine, data-driven and cross-validated models of staging are excluded by the Western status quo psychiatrists.

### The ROI score and BD

Importantly, we computed ROI in the combined study group of MDD and BD patients and observed that the ROI concept was more predictive of the phenome and present suicidal behaviors than the BD diagnosis. This can be explained by the fact that ROI quantifies the number of (hypo)manic episodes, which indicate BD but contains more information on recurrence frequency. In addition, as we have already discussed, the fact that the number of depressed and manic episodes belong to the same latent construct demonstrates that they are manifestations of the same core, which drives these manifestations. This may be explained by the fact that all individuals with mood disorders suffer from MDD or MDE, whereas only a portion of them have BD. As a result, it is puzzling that the DSM/ICS and the majority, if not all, psychiatrists classify MDD/MDE patients with manic episodes in a separate BD class and regard BD as a distinct illness, despite the fact that depressive episodes are the primary phenomenon in all mood disorder patients and (hypo)manic episodes superimpose on a depressive background. In this light, it is intriguing to note that anxiety disorders (according to DSM-IV, namely OCD, PTSD, panic disorder, and phobias) share the same phenome score as ROI and clinical ROI features, implying that when anxiety disorders and BD coexist more severe phenotypes (including MDMA and phase 3 lifetime trajectory) are shaped.

Most importantly, our MGA-PLS results show that our PLS model is not significantly different between unipolar depression and BD since none of the paths between the major disease components was significantly different between the two diagnoses. Since there are no differences between BD and unipolar depression, and ROI is a far greater predictor than BD, it appears prudent to abolish diagnoses like unipolar MDD and BD (Maes, 2022; Maes and Stoyanov, 2022). This will significantly improve mood disorder diagnosis, as in any case patients with BD are commonly misclassified as having MDD, or are underdiagnosed or overdiagnosed (Maes, 2022). This is just another example of DSM/ICD and clinicians being fixated on transforming incorrect top-down generated man-made higher-order constructs based on descriptive narratives into an incorrect gold-standard diagnosis.

### The lifetime trajectory score

We developed a simple lifetime trajectory algorithm that reflects the specific lifetime epochs of patients with mood disorders, from ACEs to ROI to phenome, including comorbid anxiety disorders that, as BD, superimpose on a depressive background characterized by ever increasing ROI. As previously stated (Maes et al., 2019), there are three phases of mood disorders: a) phase 1 (the early phase) with cognitive deficits but no significant clinical symptoms, disabilities, or changes in HR-QoL; b) phase 2 (the relapse-regression stage) with increased recurrent episodes, increased disabilities, and lower HR-QoL; and phase 3 (the suicidal-regression stage) with a very high recurrence rate of depressive episodes, highly increased suicidal behaviors, comorbidities with anxiety disorders, and a high family history and ACE scores, indicating that this phase is partly driven by increased familial and ACE load. This model includes the lifetime epochs of affective phenomenology from childhood through later phases and, therefore, this lifetime trajectory score represents different phases of retroregression and increasing impairments, as well as suicidal behaviors.

### ACEphem and family history scores

Our RADAR scores include ACEphem and a family history of mood disorders (one component extracted from depression, bipolar disorder, and suicide) as well as SUDs (one factor extracted from alcohol, TUD, and illicit drugs). Our findings indicate that ACEs mediate the effects of mood disorders and SUDs in first-degree relatives on ROI, suicidal behaviors, and the phenome, implying that interactions or cumulative effects between mental health problems in first-degree relatives and ACEs play a significant role in mood disorders. These findings demonstrate not only the effect of genetic load but also the consequences of parenteral mental health issues on mood disorders (Barnard and McKeganey, 2004; Messina and Jeter, 2012; Dakof et al., 2010). There is some evidence that mood disorders and SUD may share hereditary risk factors, and that families with SUD are more likely to have mood disordered family members than families without SUD (Quello et al., 2005).

Furthermore, mood disorders and SUDs in first-degree relatives were found to have a substantial impact on ACE in the current investigation. First-degree relatives suffering from mood disorders and SUD may become depressed, paranoid, angry, or even aggressive, creating an unstable environment for the child. Children may be terrified of a parent who repeatedly uses poor parenting tactics, such as inconsistent or harsh discipline, which can escalate to domestic violence (Stanger et al., 2004; Gershoff, 2002; Miner and Clarke-Stewart, 2008; Maes et al., 2018). Children of SUD parents are three to four times more likely to be emotionally or physically abused or neglected (Altshuler et al., 2011; McGlade et al., 2009), and parenteral alcohol use may increase the risk of poor parenting behaviors leading to a negative family environment, whereas parenteral illicit drug use increases the risk of child abuse and neglect (Kelley et al., 2010). SUDs can result in poor parenting skills and responsibilities, such as failing to provide the child’s most basic requirements for food, hygiene, safety, and stability (Barnard and McKeganey, 2004).

ACEs, particularly those accumulated during childhood, have been found to increase the risk of developing bipolar disorder and more severe phenotypes of depression later in life (Aas et al., 2016; Alvarez et al., 2022; Etain et al., 2010; Maes et al., 2021a; 2022b). Previous research has linked ACEs to recurrence of episodes (Agnew-Blais and Danese, 2016; Maes et al., 2022b), increased severity of illness, and more suicidal conduct as well as concomitant anxiety (review: Maes et al., 2021a). In is important to note that ACEs are strongly associated with increased immunoreactivity in the cytokine network as well as increased growth factor responsivity, indicating that the this connection may cause increased ROI and phenome scores (Maes et al., 2022b).

### Limitations

One limitation of this study is the drug state of the patients, which could impact the scores of the current phenome, thereby making the estimates less precise. Indeed, the phenome2 score was positively associated with the use of antidepresants, antipsychotics and mood stabilizers but not lithium, although after controlling for ACEphem and ROI, these treatment effects were no longer significant. Moreover, ACEphem and ROI2 were positively associated with the current use of antidepressants, antipsychotics, lithium, and mood stabilizers, indicating that both ACE and ROI at least in part determine the use of psychotropic drugs. It follows also that psychotropic drugs have no effects on the course and lifetime trajectory of patients with mood disorders, and thus are inadequate in treating mood disorders or preventing recurrent episodes. Second, because our phenome scores were based on a cohort of patients in (partial) remission, future research should recalculate the score rules for individuals in the acute episode, including the severity of (hypo)mania. Third, the current study was conducted on a Brazilian population and so may not be generalizable to other nations or ethnicities, but the findings have recently been replicated in a Thai group (Maes et al., TBS). Fourth, it may be argued that the relatively small sample size makes the regression analysis’ parameter estimates less precise. However, increasing the number of participants would result in an increase in analytical error when assessing biomarkers, particularly in the lower analyte concentration ranges, due to increased inter-assay (and inter-plate) variation, which might dramatically impair the model’s overall precision (Maes et al., 1994). The current study’s post-hoc minimum sample size estimation suggests that 173 people are required to have a power of 0.9 at p=0.05. As a result, we urge that future biomarker research (except for genetic studies), particularly when using neuro-immune tests, be carried out on a study sample of maximal 180 to reduce analytical error. Researchers frequently overlook the fact that as analytical error increases, it might exceed between-subject variation, resulting in uninterpretable data.

## Conclusions

In this study, we explained how to compute RADAR scores for diagnosing MDMD and scoring ROI, lifetime (LT) and current suicidal SI and SA, the phenome of mood disorders, and the lifetime trajectory of mood disorder patients (extending from a family history of mood disorders and substance abuse to adverse childhood experiences, ROI, and the phenome). In addition, we show how to plot 12 key mood disorder features in a single RADAR graph that displays all features in a two-dimensional plot. Those graphs allow us to display the characteristics of a specific patient as an idiomatic fingerprint and to estimate the key features and severity of illness at a glance. The RADAR scores, and not the incorrect DSM/ICD diagnosis or inaccurate folk-psychology derived rating scales, should be employed in biomarker research. The findings reinforce our view that the pathophysiology underlying ROI, as well as the impact of ACEs on ROI (immune and growth factor responsiveness, as well as decreased antioxidant defenses), are the most important drug targets for treating and preventing mood disorders. Pan-omics data should be used to create pathway phenotypes (e.g. ROI-biomarker phenotype) that should be included in the RADAR graph, as well as to reify the diagnostic case definitions of MDMD/SDMD as endophenotype classes, and to demarcate first-episode patients who will develop MDMD from people with a distress response, including grief and demoralization.

## Declarations

### Ethics approval and consent to participate

Approval for the study was obtained from the Institutional Review Board of the State University of Londrina, Brazil (protocol number: CAAE 34935814.2.0000.5231). All participants gave written informed consent prior to taking part in the study.

### Availability of data and materials

The dataset (PLS models) generated during and/or analyzed during the current study will be available from Prof. Dr. Michael Maes upon reasonable request and once the dataset has been fully exploited by the authors.

## Acknowledgements

We gratefully acknowledge the help of all psychiatry staff involved in the execution of this study.

## ELECTRONIC SUPLEMENTARY FILE (ESF)

**ESF, Figure 1.**
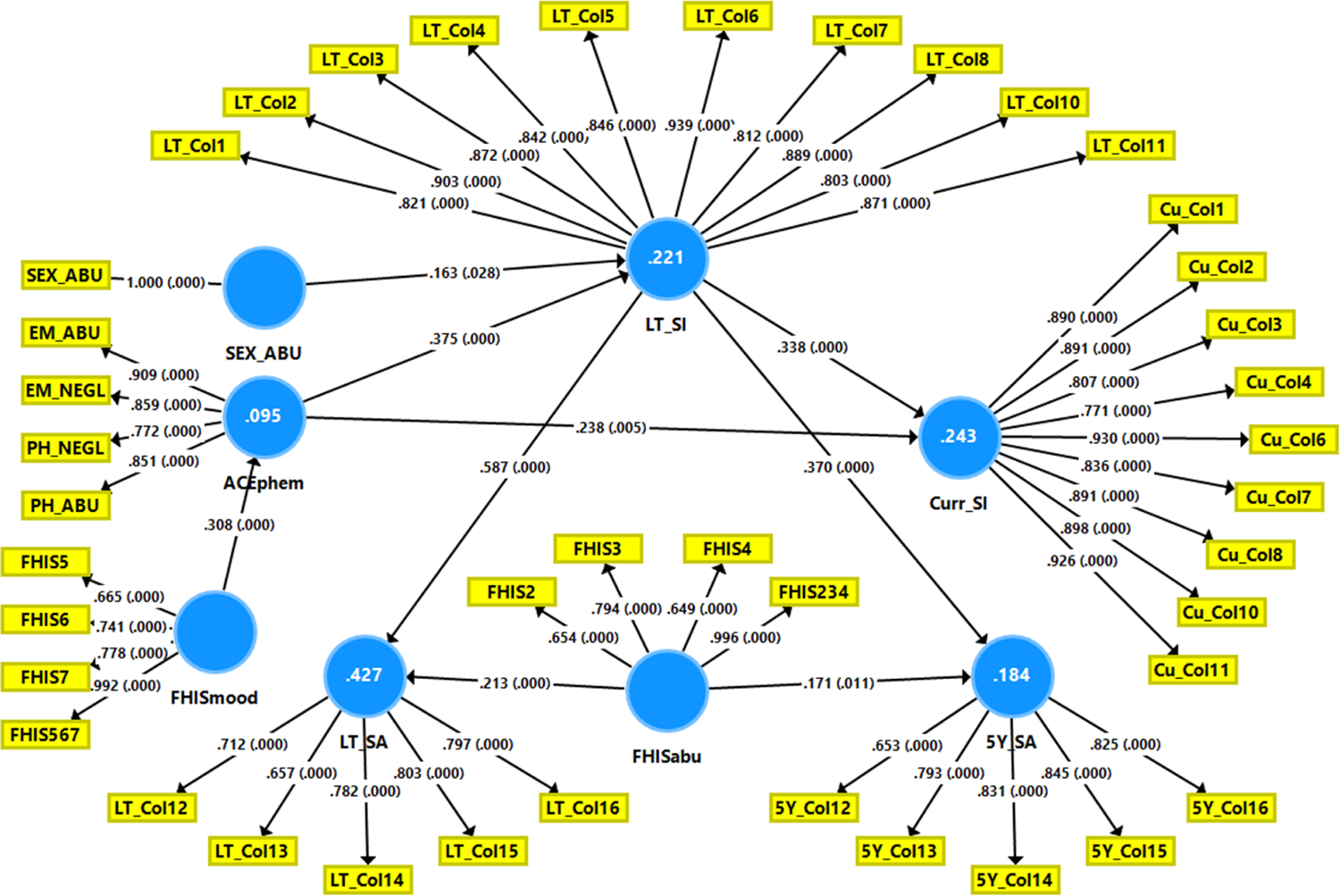
Partial Least Squares (PLS) model of suicidal behaviors in mood disorders. The final outcome variables are current suicidal ideation (curr SI) and suicidal attempts, last 5 years (5Y SA). Predictors are adverse childhood experiences (ACE) including physical and emotional abuse (PH_ABU and EM_ABU) and physical and emotional neglect (PH_NEGL and EM_NEGL) (combined into ACEphem) and sexual abuse; a family history (FHIS) of depression (FHIS5), bipolar disorder (FHIS6) and suicide attempts (FHIS7) (combined into FHISmood) and FHIS of tobacco use disorder (FHIS2), alcohol use disorder (FHIS3), substance use disorders (illicit drugs: FHIS4) (combined into FHISabu). The effects of ACEs and HFIS on Curr SI and 5Y SA are mediated via lifetime (LT) SI and SA. All indicators are entered as latent vectors, except sexual abuse. Only significant paths are shown. Displayed are path coefficients (with p values) and factor loadings (with p values). Figures in blue circles indicate explained variance.

**ESF, Figure 2.**
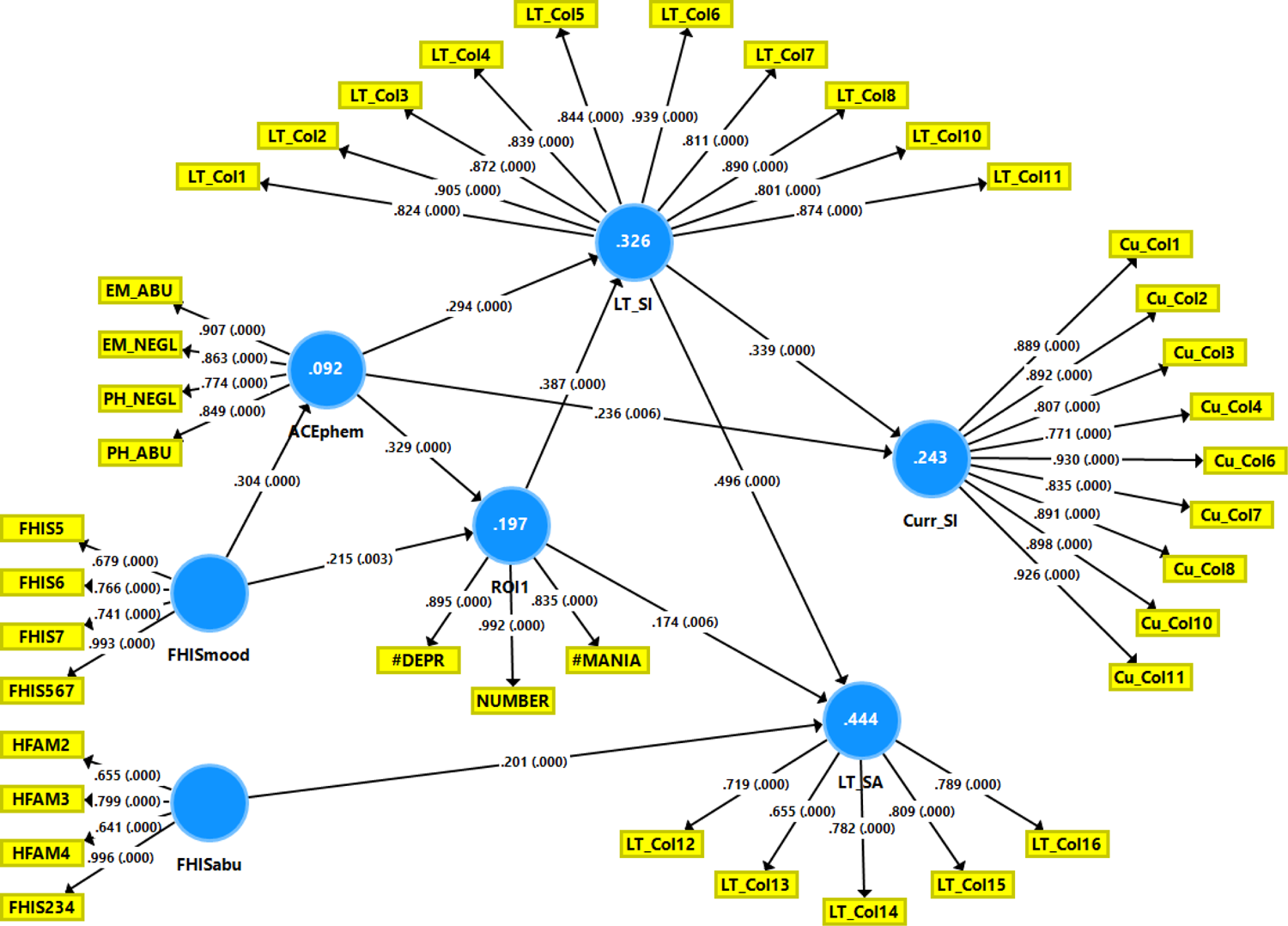
Partial Least Squares (PLS) model of suicidal behaviors and recurrence of illness (ROI) in mood disorders. The final outcome variable is current suicidal ideation (curr SI). Predictors are adverse childhood experiences (ACE) including physical and emotional abuse (PH_ABU and EM_ABU) and physical and emotional neglect (PH_NEGL and EM_NEGL) (combined into ACEphem), and a family history (FHIS) of depression (FHIS5), bipolar disorder (FHIS6) and suicide attempts (FHIS7) (combined into FHISmood) and FHIS of tobacco use disorder (FHIS2), alcohol use disorder (FHIS3), substance use disorders (illicit drugs: FHIS4) (combined into FHISabu). The effects of ACEs and HFIS on Curr SI are mediated via lifetime (LT) SI and LT SA and ROI, conceptualized as a factor extracted from number of depression (#depr) and (hypo)mania (#mania) episodes and total number of episodes. All indicators are entered as latent vectors extracted from the relevant indicators. Only significant paths are shown. Displayed are path coefficients (with p values) and factor loadings (with p values). Figures in blue circles indicate explained variance.

**ESF, Figure 3.**
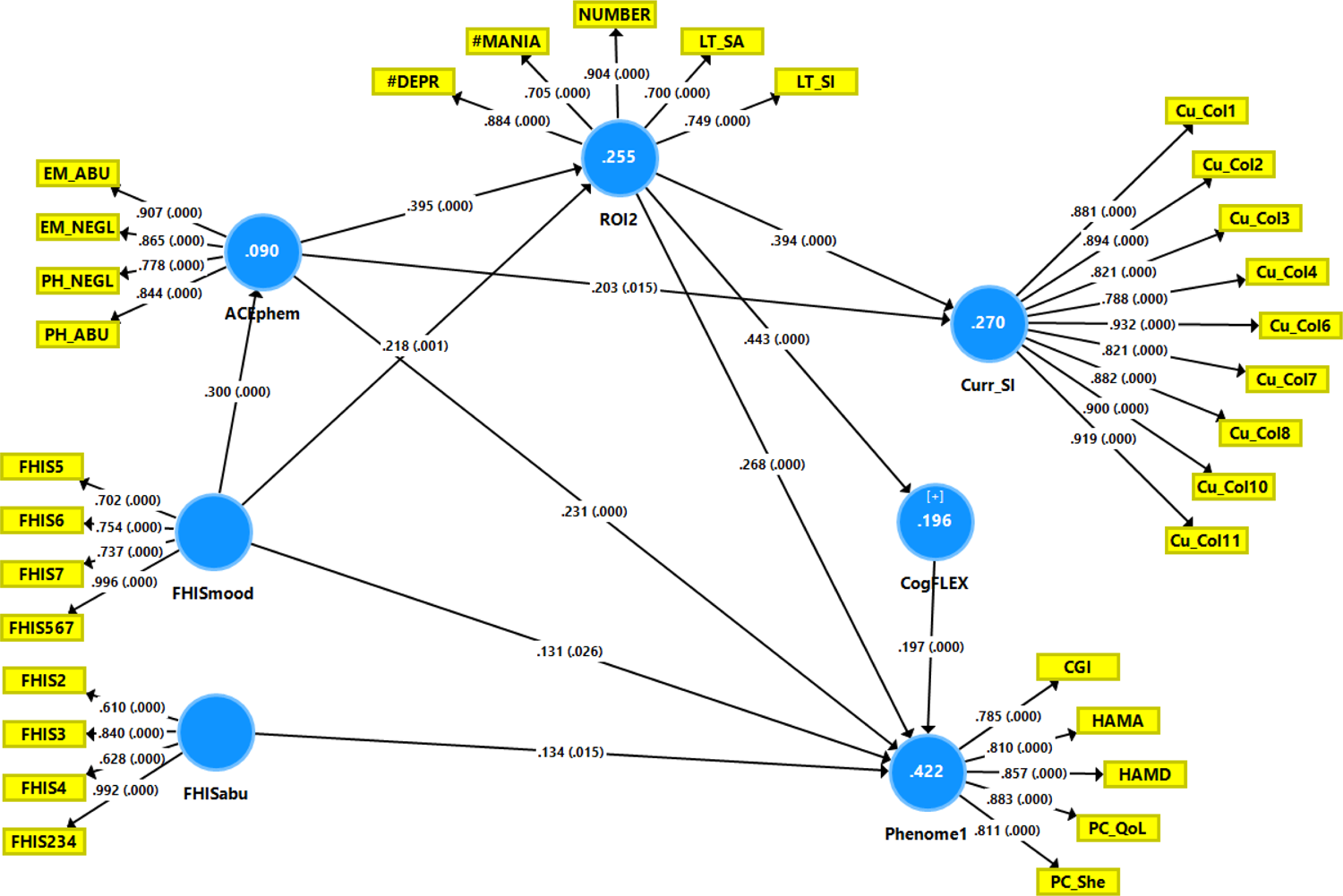
Partial Least Squares (PLS) model of suicidal behaviors, a comprehensive recurrence of illness (ROI2) index, cognitive deficits (CogFLEX) and the phenome of mood disorders. The final outcome variables are current suicidal ideation (curr SI) and the phenome of mood disorders, conceptualized as a factor extracted from the phenome and phenomenome, comprising the Hamilton Depression and Anxiety Rating Scale (HAMD/HAMA) scores; four quality of life domains (PC_QoL); five disability Sheehan (PC_She..) scores; and the Clinical Global impression (CGI) score. Predictors are adverse childhood experiences (ACE) including physical and emotional abuse (PH_ABU and EM_ABU) and physical and emotional neglect (PH_NEGL and EM_NEGL) (combined into ACEphem), and a family history (FHIS) of depression (FHIS5), bipolar disorder (FHIS6) and suicide attempts (FHIS7) (combined into FHISmood) and FHIS of tobacco use disorder (FHIS2), alcohol use disorder (FHIS3), substance use disorders (illicit drugs: FHIS4) (combined into FHISabu). The effects of ACEs and HFIS on Curr SI and the phenome are mediated via ROI conceptualized as a factor extracted from lifetime (LT) SI and LT SA, number of depression (#depr) and (hypo)mania (#mania) episodes and total number of episodes, and CogFLEX, namely cognitive deficits in verbal fluency and executive functions. All indicators are entered as latent vectors extracted from the relevant indicators, except CogFLEX which is entered as a single indicator. Only significant paths are shown. Displayed are path coefficients (with p values) and factor loadings (with p values). Figures in blue circles indicate explained variance.

**ESF Figure 4.**
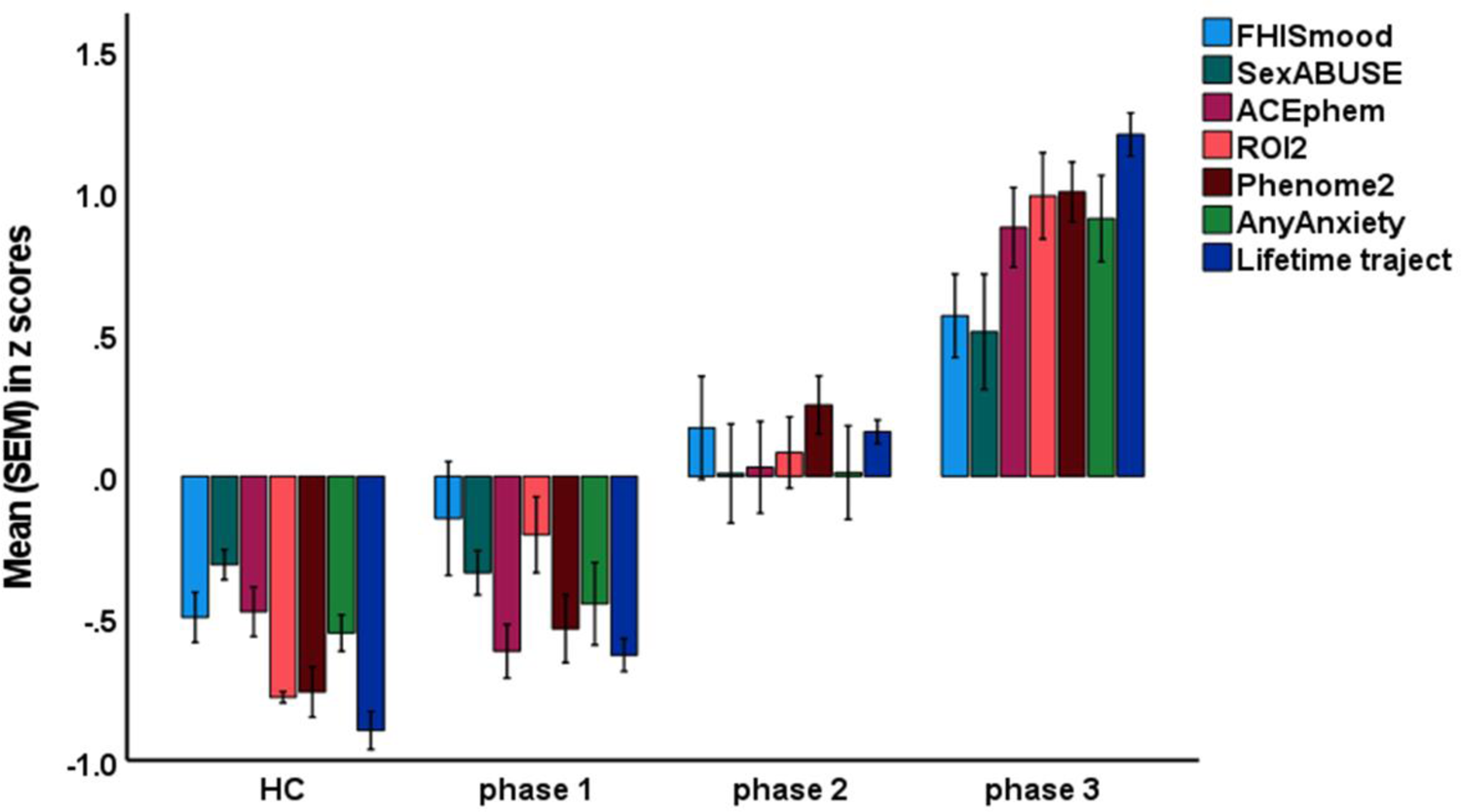
Clustered bar graph showing the RADAR score differences between healthy controls (HC) and patients with mood disorders divided into three lifetime trajectory phases. FHISmood: a family history of mood disorders and suicide; ACEphem: adverse childhood experiences (neglect and abuse); ROI2: recurrence of illness index; phenome2: an index of phenome and phenomenome; any anxiety: number of comorbid anxiety disorders.

---

**ESF Table 1.**
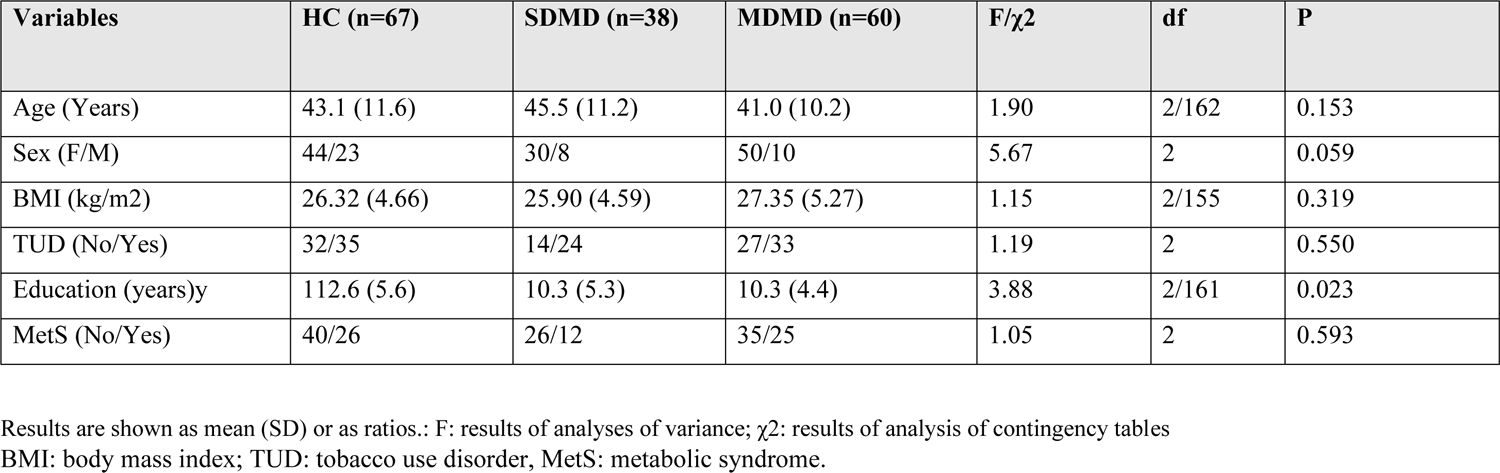
Demographic data of the healthy controls (HC) and mood disorder patients divided into those with major dysmood disorder (MDMD) and simple dysmood disorder (SDMD).

**ESF, Table 2.** Results of multiple regression analysis with ROI2 INDEX as dependent variable.

|  |  | Bootstrap for Coefficients |  |  |  |  |  |
| --- | --- | --- | --- | --- | --- | --- | --- |
|  |  |  |  |  | Bootstrap<sup>a</sup> | 95% Confidence Interval |  |
| Model |  | B | Bias | Std. Error | Sig. (2-tailed) | Lower | Upper |
| 1 | Constant | -.803 | .001 | .011 | <.001 | -.823 | -.779 |
|  | Total number episodes | .088 | .000 | .002 | <.001 | .084 | .093 |
|  | C-SSRS-6 LT | .139 | -.002 | .010 | <.001 | .115 | .157 |
|  | C-SSRS-12 LT | .082 | .008 | .025 | .002 | .067 | .163 |

a. Bootstrap results are based on 1000 bootstrap samples
C-SSRS-6-LT: what is the most intense type of ideation during life
C-SSRS-12 LT: number of suicidal attempts, lifetime

**ESF, Table 3.** Results of multiple regression analysis with lifetime suicidal behaviors as dependent variable.

|  |  | Bootstrap for Coefficients |  |  |  |  |  |
| --- | --- | --- | --- | --- | --- | --- | --- |
|  |  | B | Bias | Std.<br>Error | Bootstrap<sup>a</sup> |  |  |
| Model |  |  |  |  | Sig. (2-tailed) | 95% Confidence<br>Interval |  |
|  |  |  |  |  |  | Lower | Upper |
| 1 | (Constant) | -.722 | -.001 | .011 | <.001 | -.744 | -.700 |
|  | C-SSRS-2-LT | .268 | .004 | .084 | .006 | .095 | .425 |
|  | C-SSRS-4 LT | .340 | -.014 | .129 | .014 | .084 | .586 |
|  | C-SSRS-5 LT | .432 | -.012 | .150 | .006 | .121 | .673 |
|  | C-SSRS-6 LT | .121 | -.004 | .041 | .011 | .041 | .205 |
|  | C-SSRS-7 LT | .073 | .004 | .038 | .076 | -.001 | .149 |
|  | C-SSRS-12 LT | .172 | .023 | .064 | .003 | .128 | .372 |

a. Unless otherwise noted, bootstrap results are based on 1000 bootstrap samples
C-SSRS-2-LT: death wish during life
C-SSRS-4 LT: active suicidal ideation without specific plan, lifetime
C-SSRS-5-LT: active suicidal ideation with specific plan, lifetime
C-SSRS-6-LT: what is the most intense type of ideation during life
C-SSRS-7 LT: frequency of suicidal ideation, lifetime
C-SSRS-12 LT: number of suicidal attempts, lifetime

**ESF, Table 4.** Results of multiple regression analysis with current suicidal ideation as dependent variable.

|  |  | Bootstrap for Coefficients |  |  |  |  |  |
| --- | --- | --- | --- | --- | --- | --- | --- |
|  |  |  |  |  | Bootstrap<sup>a</sup> | 95% Confidence Interval |  |
| Model |  | B | Bias | Std. Error | Sig. (2-tailed) | Lower | Upper |
| 1 | (Constant) | -.414 | -5.551E-16 | 2.843E-16 | <.001 | -.414 | -.414 |
|  | C-SSRS-1 Current | 1.013 | -.011 | .132 | <.001 | .730 | 1.256 |
|  | C-SSRS-3 Current | .979 | -.007 | .282 | .006 | .435 | 1.558 |
|  | C-SSRS-6 Current | .457 | .006 | .090 | <.001 | .298 | .643 |

a. Unless otherwise noted, bootstrap results are based on 1000 bootstrap samples
C-SSRS-1-current: wish to be death, current
C-SSRS-3 current: active suicidal ideation with any method, current
C-SSRS-6-current: what is the most intense type of ideation, current

**ESF, Table 5.** Results of multiple regression analysis with phenome2 as dependent variable.

|  |  | Bootstrap for Coefficients |  |  |  |  |  |
| --- | --- | --- | --- | --- | --- | --- | --- |
|  |  |  |  |  | Bootstrap<sup>a</sup> | 95% Confidence Interval |  |
| Model |  | B | Bias | Std. Error | Sig. (2-tailed) | Lower | Upper |
| 1 | (Constant) | -1.523 | -.009 | .053 | <.001 | -1.641 | -1.429 |
|  | C-SSRS-1 Current | .363 | -.002 | .090 | <.001 | .187 | .536 |
|  | C-SSRS-4 Current | .387 | -.002 | .123 | .002 | .141 | .616 |
|  | Total HAMD | .055 | .000 | .006 | <.001 | .043 | .068 |
|  | Total HAMAA | .030 | .000 | .004 | <.001 | .020 | .038 |
|  | CGI | .200 | .003 | .024 | <.001 | .160 | .252 |

a. Unless otherwise noted, bootstrap results are based on 1000 bootstrap samples
C-SSRS-1-current: wish to be death, current
C-SSRS-4 current: active suicidal ideation without any specific plan, current
HAMD/HAMA: the Hamilton Depression and Anxiety Rating Scale scores
CGI: Clinical Global impression score

**ESF, Table 6.**
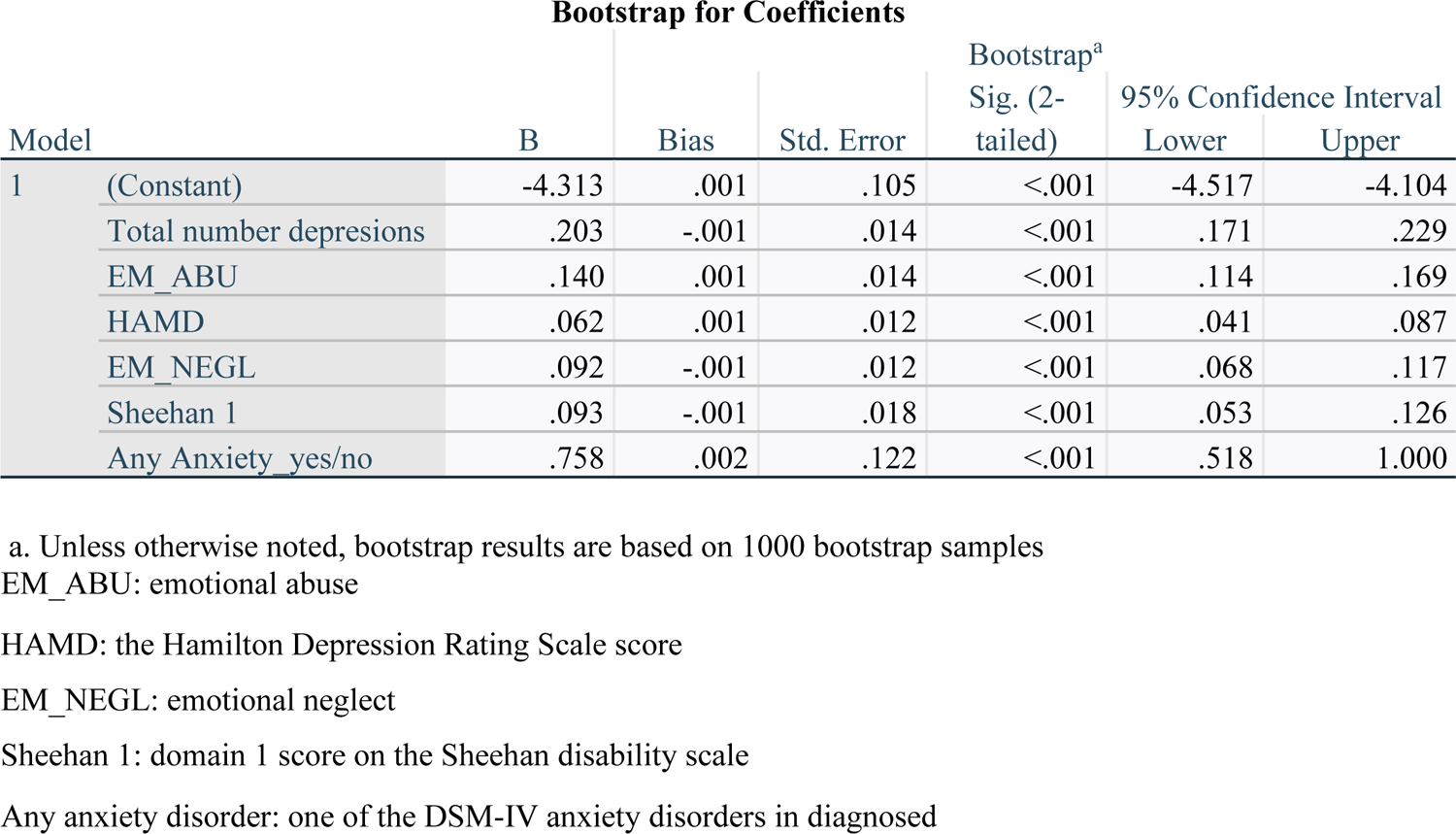
Results of multiple regression analysis with the lifetime trajectory score as dependent variable.

|  |  | Bootstrap for Coefficients |  |  | Bootstrap<sup>a</sup> |  |  |
| --- | --- | --- | --- | --- | --- | --- | --- |
| Model |  | B | Bias | Std. Error | Sig. (2-tailed) | 95% Confidence Interval |  |
|  |  |  |  |  |  | Lower | Upper |
| 1 | (Constant) | -4.313 | .001 | .105 | <.001 | -4.517 | -4.104 |
|  | Total number depresions | .203 | -.001 | .014 | <.001 | .171 | .229 |
|  | EM_ABU | .140 | .001 | .014 | <.001 | .114 | .169 |
|  | HAMD | .062 | .001 | .012 | <.001 | .041 | .087 |
|  | EM_NEGL | .092 | -.001 | .012 | <.001 | .068 | .117 |
|  | Sheehan 1 | .093 | -.001 | .018 | <.001 | .053 | .126 |
|  | Any Anxiety_yes/no | .758 | .002 | .122 | <.001 | .518 | 1.000 |

a. Unless otherwise noted, bootstrap results are based on 1000 bootstrap samples
EM\_ABU: emotional abuse
HAMD: the Hamilton Depression Rating Scale score
EM\_NEGL: emotional neglect
Sheehan 1: domain 1 score on the Sheehan disability scale
Any anxiety disorder: one of the DSM-IV anxiety disorders in diagnosed

**ESF 2, Table 7.** Relevant cut-off values for the key standardized predicted values of the key features of mood disorders

| Predicted values | Q50 | Q75 |
| --- | --- | --- |
| 1. Recurrence of illness index | 0.197 | 0.914 |
| 2. Lifetime suicidal behaviors | 0.153 | 0.1252 |
| 3. Current suicidal ideation | 1.1025 | 1.895 |
| 4. Phenome2 | 0.289 | 0.946 |
| 5. Life trajectory | 0.392 | 0.959 |

Q50/Q75 are computed on the values obtained in patients.

**ESF, Table 8.** Point-biserial correlations between the drug state variables and a family history of mood disorders and suicidal attempts (FHISmood), adverse childhood experiences of physical and emotional abuse and neglect (ACEphem), recurrence of illness (ROI2) and the phenome-phenomenome index (phenome2).

Correlations
|  |  | antidep. | Antipsy | lithium | moodst |
| --- | --- | --- | --- | --- | --- |
| FHISmood | Pearson Correlation | .203** | .208** | .311** | .268** |
|  | Sig. (2-tailed) | 0.010 | 0.008 | 0.000 | 0.001 |
|  | N | 162 | 162 | 161 | 161 |
| ACEphem | Pearson Correlation | 0.118 | .234** | .258** | .306** |
|  | Sig. (2-tailed) | 0.135 | 0.003 | 0.001 | 0.000 |
|  | N | 162 | 162 | 161 | 161 |
| ROI2 | Pearson Correlation | .276** | .363** | .412** | .438** |
|  | Sig. (2-tailed) | 0.000 | 0.000 | 0.000 | 0.000 |
|  | N | 162 | 162 | 161 | 161 |
| Phenome2 | Pearson Correlation | .290** | .267** | 0.120 | .308** |
|  | Sig. (2-tailed) | 0.000 | 0.001 | 0.128 | 0.000 |
|  | N | 162 | 162 | 161 | 161 |

\*. Correlation is significant at the 0.05 level (2-tailed).
\*\*. Correlation is significant at the 0.01 level (2-tailed).
Antidep: antidepressants; antipsy: antipsychotics, moodst: mood stabilizers.

